# PanoraOnc: A pan-cancer clinico-genomic AI model for transferable outcome predictions

**DOI:** 10.64898/2026.08.17.26354679

**Authors:** Manuel Schürch, Jacob Geisberg, Cameron T. Flower, Ayyuce Begum Bektas, Thomas O. McDonald, Satyakam Mishra, Christopher Graser, Jennifer Altreuter, Guruprasad Ananda, Genevieve Boland, David Liu, Kenneth L. Kehl, Franziska Michor

**Author notes:** These authors contributed equally (Jacob Geisberg, Cameron T. Flower, Ayyuce Begum Bektas).

## Abstract

Progress in precision oncology, including biomarker discovery and individualized treatment selection, is limited by the complexity of clinico-genomic data and the scarcity of large multimodal patient cohorts. Here, we introduce *PanoraOnc*, a pan-cancer artificial intelligence (AI) model pretrained on real-world clinical, genomic, and imaging data from 84,131 patients spanning 66 cancer types. *PanoraOnc* enables transferable treatment outcome prediction through pan-cancer pretraining and generalizes to unseen cohorts across cancer types, institutions, and therapeutic settings. Evaluation and fine-tuning were performed on cohorts comprising diverse modalities, including clinical features, targeted gene panels, immunofluorescence imaging, whole-exome sequencing, and transcriptomic profiles. Across these settings, *PanoraOnc* consistently outperforms statistical, machine-learning, survival, and AI baselines, with the largest improvements observed in zero- and few-shot scenarios, demonstrating that large-scale clinico-genomic pretraining enables robust and generalizable outcome predictions across previously unseen conditions. In addition, *PanoraOnc* supports biomarker discovery through explainable AI, revealing both established and underappreciated features, including tumor-infiltrating clonal hematopoiesis, oncogenic signaling pathways, and DNA damage response mechanisms in immunotherapy-treated melanoma and non-small cell lung cancer. Furthermore, *PanoraOnc* enables the identification of patient subgroups potentially benefitting from alternative treatments by estimating personalized treatment outcomes across therapeutic scenarios. These findings establish pan-cancer multimodal pretraining as a scalable paradigm for AI-assisted discovery in precision oncology.

## Introduction

Personalized therapeutic decision-making in oncology remains a major challenge due to the heterogeneity of demographic, clinical, genomic, transcriptomic, and imaging data across patients, cancer types, and evolving treatment policies^1,2^. Despite substantial progress, many predictive models remain constrained by small cohort sizes, reliance on single-modality data, limited generalization beyond the institutions in which they are developed, and insufficient capacity to link the complex, nonlinear relationships between multimodal patient profiles, therapeutic interventions, and clinical outcomes, with few addressing actionable treatment decisions^2–4^. Consequently, model performance often degrades when applied to data from new or unseen cohorts, treatment regimes, or clinical settings^3,5,6^. Addressing these limitations requires robust zero-shot generalization for counterfactual treatment outcome prediction — the ability to make reliable and transferable predictions for previously unseen patients, cancers, therapies, and institutions without access to closely matched training cohorts.

Current artificial intelligence (AI) approaches offer a promising avenue to meet this challenge and have shown substantial promise across a wide range of oncology applications, including cancer diagnosis and classification^7^, electronic health record representation learning and outcome prediction^8–12^, and the development of radiologic^13^ and histopathologic^14,15^ foundation models. Despite this progress, a central unmet challenge remains learning transferable treatment–response representations from real-world, large-scale, high-dimensional, and multimodal clinico-genomic data. Integrating such representations into unified pretrained models is essential to support zero-and few-shot predictions for data-scarce and unseen conditions. Such capabilities are particularly valuable in complex clinical settings such as molecular tumor boards, where treatment recommendations often rely on incomplete evidence^16^, particularly for rare cancers, uncommon molecular alterations, late-stage disease, off-label use, or data-scarce early-stage clinical trials. AI systems with these capabilities could enable researchers to systematically explore patient-specific treatment outcome scenarios and derive mechanistic insights from integrated multimodal data.

To address these issues, we introduce *PanoraOnc*, a pan-cancer clinico-genomic AI framework for predicting cancer therapy outcomes (Fig. 1A), pretrained on multimodal data from heterogenous datasets and treatment regimens (N = 84,131, Fig. 1B). These datasets encompass multimodal patient data (Fig. 1C), including electronic health records (demographics, treatments, and outcomes), targeted gene panels (somatic mutations and copy number alterations), whole-exome sequencing (WES), RNA sequencing (RNA-seq) data, and immune-profiled imaging data (IP). *PanoraOnc* generates probabilistic outcome predictions for treatment response and one- to three-year survival alongside in-context biomarker identification and analysis of hypothetical treatment scenarios (Fig. 1D, Methods). *PanoraOnc*’s pan-cancer pretraining enables transfer learning across cancer types, modalities, and therapeutic contexts, laying the foundation for a unified AI framework in oncology.

**Figure 1:**
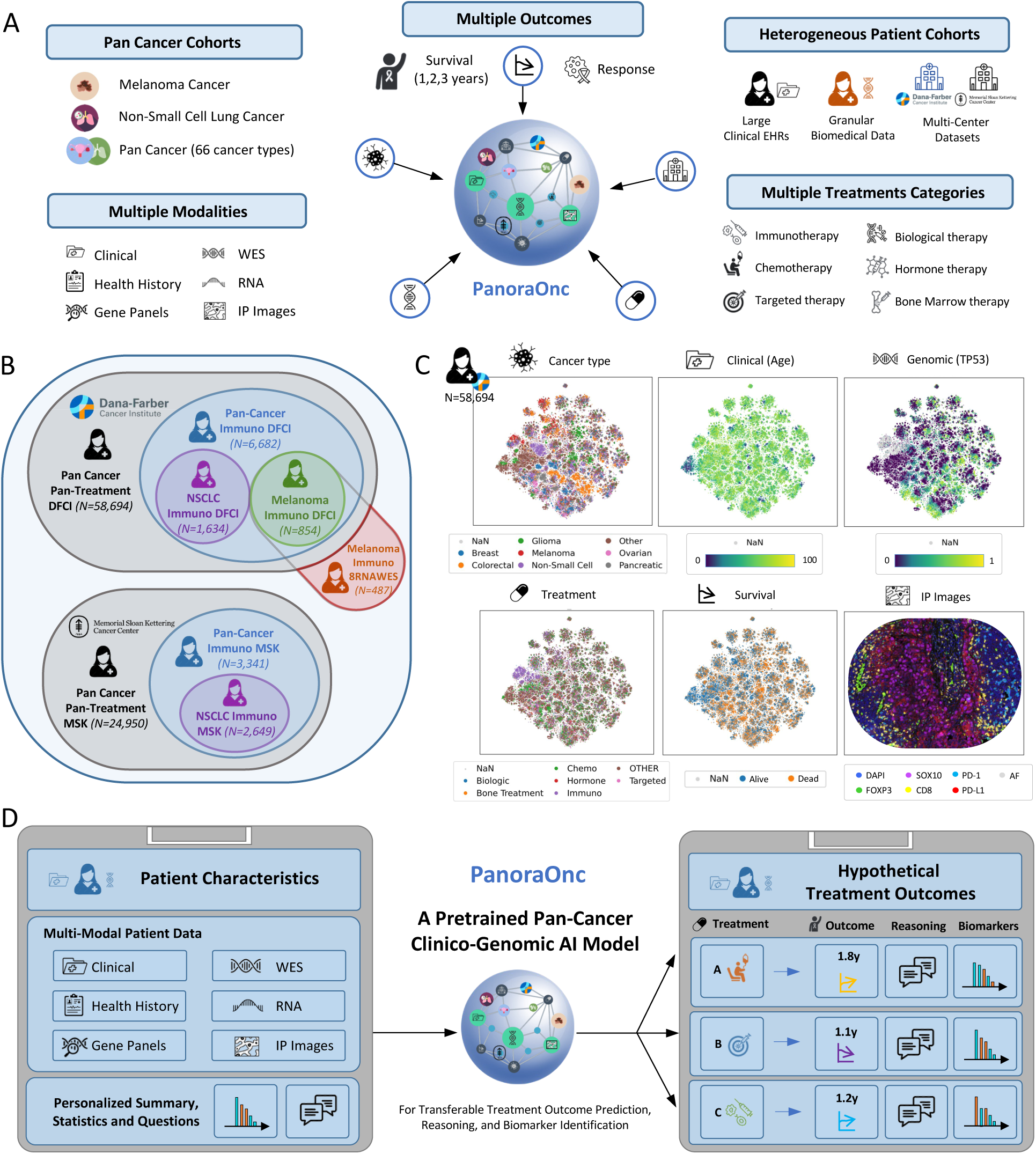
Development of PanoraOnc, a Pretrained Pan-Cancer Clinico-Genomic AI Model. **(A) Goal:** Development of a pretrained pan-cancer clinico-genomic AI model for quantitative and qualitative prediction of treatment outcomes by integrating multiple data modalities, clinical endpoints, cancer types, and heterogenous patient cohorts. **(B) Cohorts:** Integration of heterogeneous clinico-genomic datasets with overlapping modalities at DFCI (N=58,694), MSK (N=24,950), and eight granular cohorts (8RNAWES) with a particular focus on non-small cell lung cancer (NSCLC) and melanoma patients treated with immunotherapy. **(C) Data:** Large-scale pan-cancer real-world patient data encompassing clinical, genomic, treatment, imaging, and outcome modalities. **(D) Model:** Based on clinical and biomedical multimodal patient characteristics, PanoraOnc generates quantitative and qualitative predictions of response to different treatment options, along with reasoning and biomarker identification.

### Large-scale real-world multimodal clinico-genomic cancer patient cohorts

In 2011, Dana-Farber Cancer Institute (DFCI) launched *Profile*^17^, one of the most comprehensive personalized cancer medicine initiatives in the US, with over 100,000 patients consented for the collection of clinico-genomic information, including information on treatments, cancer type, stage, progression, and survival, as well as *Profile*^18^ panel genomic information. The *Pan-Cancer Pan-Treatment DFCI* cohort comprises N = 58,694 patients (Fig. 2A, Fig. S1, Table S1) and 66 cancer types with diverse treatment categories (immunotherapy, chemotherapy, targeted therapy, biological therapy, hormone therapy, bone therapy) defined as the first applied treatment category after genomic sequencing. We defined several cohort subsets (Fig. 1B and 2A), including the *Pan-Cancer Immuno DFCI* cohort (N = 6,682, Fig. S2, Table S2), the *non-small cell lung cancer* (*NSCLC) Immuno DFCI* (N = 1,634, Fig. S3, Table S3), and the *Melanoma Immuno DFCI* (N = 854, Fig. S4, Table S4) cohorts. The *Pan-Cancer Imaging DFCI* cohort consists of the immune-profiled Immunoprofile (IP) cohort^19^ (N=1,499) for whom spatial single-cell resolution immunofluorescence data (DAPI, cytokeratin/SOX10/PAX8, FOXP3, CD8, PD-1, PD-L1, autofluorescence) are available. We also included an independent cohort from Memorial Sloan Kettering Cancer Center (MSK), *Pan-Cancer Pan-Treatment MSK*^20^, with a total of N = 24,950 patients and 5 cancer types (Fig. 2A, Fig. S5, Table S5), including the *Pan-Cancer Immuno MSK* (N = 3,341, Table S6) and the *NSCLC Immuno MSK* (N = 2,649, Table S7) cohorts. In addition, we incorporated the *8RNAWES* cohort (N = 487, Fig. S6, Table S8) based on eight international melanoma patient cohorts, including five datasets^21–25^ with both WES and RNAseq (N = 347) and three cohorts^26–28^ with RNAseq only (N = 140). These real-world cohorts are highly heterogeneous, spanning a wide range of treatment regimens, outcome distributions, cohort shifts, cancer types, biopsy sites, different missing data patterns, and potential treatment assignment biases. All clinical features and genomic features in the DFCI, MSK, and 8RNAWES cohorts were standardized and preprocessed with the same pipelines for harmonizing the input into the ML and AI models (Methods).

**Figure 2:**
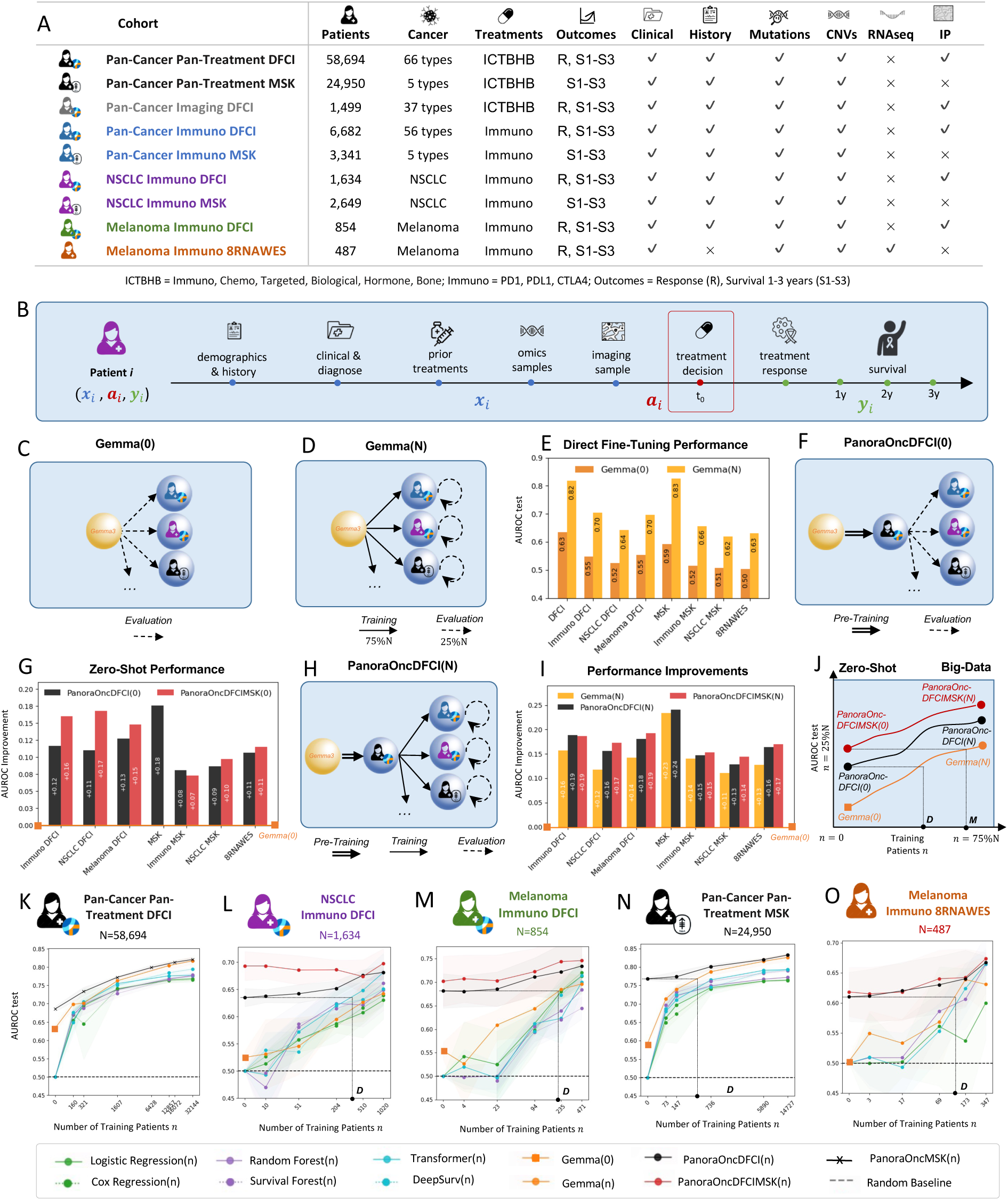
Cohorts, Models, and Performance Results of PanoraOnc. **(A)** Table of all investigated cancer cohorts showing patient numbers, treatment regimes, clinical endpoints, and available data modalities, including clinical, genomic, transcriptomic, and imaging-based (IP) data. **(B)** Schematic of longitudinal multi-model patient data representation, with a treatment decision point at t_0_. All data modalities preceding this point are used as model input, while subsequent events are treated as treatment outcomes. **(C)** Evaluation of the base model Gemma(0) on different cohorts. **(D)** Fine-tuned Gemma(N) model on individual cohorts and evaluation on held-out test sets. **(E)** Direct fine-tuning performance results for Gemma(N) compared to zero-shot inference of base model Gemma(0) for 2-year survival outcome prediction. **(F)** Pretrained PanoraOncDFCI(0) model on the large pan-cancer DFCI cohort with zero-shot evaluations on internal and external cohorts. **(G)** Zero-shot performance improvements of PanoraOncDFCI(0) and PanoraOncDFCIMSK(0) over the base model Gemma(0) after pan-cancer pre-training on the DFCI cohort alone or both the DFCI and MSK cohorts without using any cohort-specific data. **(H)** Fine-tuned PanoraOncDFCI(N) model starting from the pretrained model. **(I)** Performance improvements over the base model of the directly fine-tuned Gemma(N), pretrained and fined-tuned PanoraOncDFCI(N) and PanoraOncDFCIMSK(N) models in the asymptotic case. **(J)** Schematic overview of the different models as a function of training cohort size n, ranging from zero-shot to asymptotic settings. The crossover points D and M indicate situations in which the best directly trained baseline performs better than zero-shot PanoraOncDFCI(0) or PanoraOncDFCIMSK(0). **(K-M)** Detailed performance results of PanoraOnc models compared to ML and AI baselines for the internal DFCI cohorts and **(N-O)** external evaluations on the MSK and 8RNAWES cohorts. Additional results for all outcomes and cohorts are provided in Figure 3 and S7.

### Development of the pan-cancer clinico-genomic multimodal *PanoraOnc* model

*PanoraOnc* is built on an open-weight, medium-sized vision-language model (VLM), the Gemma3-12B model^29^, and it was trained on diverse multimodal cancer datasets varying in size and modality (Fig. 2A). Each patient case is represented using text and image-derived data modalities in chronological order, including past and current treatment information at a treatment decision point t_0_ (Methods), and multiple treatment outcomes such as response and one-, two-, and three-year-survival (Fig. 2B, Figs. S9A,B). This design enables flexible and contextualized modeling of heterogeneous, structured and semi-structured multimodal patient cases, including cases with missing modalities or values. These patient cases are then used for AI predictions, fine-tuning and pre-training strategies across different settings. To assess the number of samples required to learn sparse and high-dimensional patterns from multimodal data, a particular focus was on investigating model performance as a function of training cohort size, ranging from zero-shot to progressively larger training cohorts approaching the full available dataset. All ML and AI models were trained on quantitative prediction tasks, including treatment response classification and one- to three-year post-treatment survival (Methods). For *PanoraOnc*, these multiple tasks were performed directly within the VLM without an external classification head, enabling transfer learning and unified quantitative and qualitative predictions within a single, interactive AI framework.

### Fine-tuning on real-world cancer cohorts improves quantitative outcome prediction performance

We first assessed inference-only predictions from the base model *Gemma(0)* across multiple cohorts (Fig. 2C), using no training samples and 25% of held-out test samples from each cohort across four independent train-test splits. We compared this baseline with *Gemma(N)* (Fig. 2D), a cohort-specific fine-tuned version based on the Gemma3 base model trained on 75% of the available patient samples from each individual cohort and evaluated on 25% held-out test samples from the same cohort; here N denotes the total number of patient cases in that cohort. The comparison for 2-year survival prediction between these models shows that zero-shot inference with *Gemma(0)*, relying solely on prompting, demonstrated broad medical knowledge, but its quantitative performance was modest (AUROC 0.50–0.63, Fig. 2E). In contrast, cohort-specific fine-tuning improved performance, with *Gemma(N)* achieving AUROCs of 0.62–0.83 (Fig. 2E), corresponding to improvements of +0.11–0.23 over the base model. Similar patterns were observed for the other quantitative treatment outcome predictions (Fig. S7). These results demonstrate that while zero-shot inference from the untrained model *Gemma(0)* provides limited quantitative prediction capabilities, fine-tuning on real-world cancer cohorts can substantially improve performance across diverse clinico-genomic settings, although it requires access to sufficiently large cohort-specific datasets for fine-tuning.

### Large-scale pretraining on pan-cancer cohorts enables zero-shot prediction across institutions and cancer types

We then evaluated the performance of *PanoraOnc,* an AI model pretrained on large-scale pan-cancer cohorts comprising diverse real-world patient populations. *PanoraOncDFCI(0)* was pretrained on the DFCI pan-cancer pan-treatment cohort and subsequently evaluated in a zero-shot manner on independent internal and external cohorts across cancer types, treatment regimens, and institutions (Fig. 2F). Importantly, this approach did not utilize any cohort-specific training data; for instance, the model whose performance was evaluated on the DFCI melanoma immunotherapy cohort was pretrained on all cohorts but the latter. Similarly, *PanoraOnc-DFCIMSK(0)* was pretrained on both MSK and DFCI cohorts. We then evaluated the transfer learning and zero-shot capabilities of both pretrained models (Fig. 2G), showing that these models yielded substantial improvements across both internal (DFCI) and external (MSK, 8RNAWES) datasets with AUROC zero-shot improvements over the base Gemma3 model of +0.08–0.18 for *PanoraOncDFCI(0)* and +0.07–0.17 for *PanoraOncDFCIMSK(0),* corresponding to an improvement of up to +0.06 over the DFCI-only pretrained model. Notably, external zero-shot validation of *PanoraOncDFCI(0)* on the pan-cancer pan-treatment cohort from MSK achieved an AUROC of 0.77, with +0.18 performance improvement over the base model without MSK patient data pretraining. These findings demonstrate the benefits of large-scale pretraining for improved zero-shot generalization across cancer types and institutions.

### Pre-training and fine-tuning optimizes *PanoraOnc* performance

Building on the rich prior knowledge acquired by our pretrained *PanoraOnc* models, we next investigated whether additional local data of specific cohorts could further improve performance. The combined *PanoraOncDFCI(N)* model was first pretrained on the DFCI cohort and subsequently fine-tuned on cancer- or treatment-specific cohorts to further refine individualized treatment-response patterns (Fig. 2H). Importantly, throughout, samples used for pre-training, fine-tuning, and performance evaluation were strictly non-overlapping to prevent any data leakage. When comparing the performance improvements of *Gemma(N), PanoraOncDFCI(N)* and *PanoraOncDFCIMSK(N)* fine-tuned on each cohort (Fig. 2I), we obtained an increase of +0.11-0.23 for *Gemma(N),* +0.13–0.24 for *PanoraOncDFCI(N)*, and +0.14–0.19 for *PanoraOncDFCIMSK(N)* over the base model lacking fine-tuning, demonstrating that pre-training on larger cohorts consistently improves final fine-tuning performance.

Learning sparse patterns from high-dimensional multimodal clinico-genomic patient data requires large training cohorts. To systematically characterize the performance of the *PanoraOnc* models as a function of the number of varying training samples *n*, we trained a series of models specified with *PanoraOnc(n)*, spanning zero-shot (*n=0*) to asymptotic regimes (n=75%*N* where *N* is the total cohort size), with previously discussed models appearing as special cases within this continuum (Fig. 2J). For instance, *PanoraOncDFCI(n)* represents a model that was pretrained on the DFCI pan-cancer pan-treatment cohort and fine-tuned on *n* additional cohort-specific patient samples. We further quantified the number of cohort-specific training patients required for effective local training by identifying crossover points in performance (Fig. 2J). Specifically, we defined the points D and M at which the best directly trained baseline surpasses the zero-shot performance of the pan-cancer pretrained *PanoraOncDFCI(0)* and *PanoraOncDFCIMSK(0)* models, respectively, both of which do not leverage any cohort-specific training data. These crossover points provide a practical guideline for model selection, addressing the question whether, given a multimodal cohort with *n* patients, a local model should be trained from scratch instead of using a pan-cancer–pretrained *PanoraOnc* model.

To this end, we trained and evaluated *Gemma(n)*, *PanoraOncDFCI(n)*, and *PanoraOncDFCIMSK(n)* across increasing numbers of fine-tuning samples *n*, from zero-shot (*n*=0) to asymptotic settings (*n*=75%*N*), and compared their performance to conventional statistical, machine-learning and deep learning baselines (Ridge, Random Forest, and Transformer models) and traditional survival models (Cox regression^30^, Random Survival Forest^31^, DeepSurv^32^) trained from scratch on cohorts with the same splits, training fractions, and input features (Methods). AUROC performance comparisons for 2-year survival prediction (Figs. 2K–O) demonstrated that both the few-shot and asymptotic performance of *Gemma(n)* models were only slightly higher than, or comparable to, traditional ML and survival baselines, with advantages emerging primarily in data-rich settings such as the pan-cancer and pan-treatment cohorts from DFCI (Fig. 2K) and MSK (Fig. 2N), yielding AUROC improvements of +0.05 and +0.04, respectively, over the best-performing ML baseline, the Transformer.

In contrast, the pretrained *PanoraOnc* models consistently outperformed both directly trained AI and ML baselines across zero-shot, few-shot, and asymptotic regimes on internal (Figs. 2L–M) and external cohorts (Figs. 2N–O), with particularly large gains in low data settings. For example, in the NSCLC immunotherapy cohort from DFCI (Fig. 2L), *PanoraOncDFCI* achieves a zero-shot AUROC of 0.63 for 2-year survival prediction and remains superior to directly trained models up to a training cohort size of approximately *D* = 300. Similar crossover points were observed at *D* ≈ 220 for melanoma (Fig. 2M), *D* ≈ 500 for pan-cancer MSK (Fig. 2N), and *D* ≈ 150 for RNAWES melanoma cohorts (Fig. 2O). Moreover, pretraining on both DFCI and MSK patient data further improved performance, with *PanoraOncDFCIMSK* consistently outperforming *PanoraOncDFCI* across cohorts and training regimes. Notably, the combined pretrained model *PanoraOncDFCIMSK(0)* achieves a zero-shot AUROC of 0.69 on the NSCLC cohort (Fig. 2L), representing an improvement of +0.17 over the base model and +0.06 over *PanoraOncDFCI(0).* This performance corresponds to a crossover point larger than *D* = 1,020 patients, indicating that below this cohort size, zero-shot inference with the *PanoraOncDFCIMSK(0)* model outperforms training a cohort-specific model from scratch using only NSCLC immunotherapy patients. Results for *PanoraOncMSK* (*Fig. 2K*), pretrained only on MSK and evaluated on DFCI cohorts, demonstrate further generalizability across hospital systems. In sum, our pretrained models (Fig. 2) on large-scale real-world data produce accurate quantitative predictions across multiple cohorts, with strong zero-shot and few-shot performance gains compared to non-pretrained models, establishing them as a flexible and robust foundation for transferring and generalizing treatment outcome prediction across different settings and conditions.

### PanoraOnc enables transferable multi-tasks predictions with multimodal patient data

We next evaluated *PanoraOnc* across multiple treatment outcome prediction tasks and performance metrics for the DFCI NSCLC immune cohort (Fig. 3A). The benefits of multimodal pretraining were consistently observed across one-, two-, and three-year survival predictions, as well as treatment response predictions, measured by AUROC and compared to a range of survival, ML and AI baselines (Fig. 3B-E). The biggest performance gains were observed in zero- and few-shot settings. Notably, zero-shot performance improved progressively with increasingly comprehensive pretraining, from *PanoraOncDFCIImmuno(0)*, to *PanoraOncDFCI(0)*, and *PanoraOncDFCIMSK(0)*, demonstrating the value of incorporating more patient cases (Fig. 3C). Similar improvements were observed across complementary evaluation metrics, including precision-recall area under curve (PRAUC, Fig. 3G), Brier-Scores (Fig. 3H), and calibration intercepts and slopes (Fig. 3G,H), demonstrating that pretraining improves not only discrimination but also predictive calibration and robustness across clinical endpoints. Detailed results for all cohorts are provided in Fig. S7 and S8.

**Figure 3:**
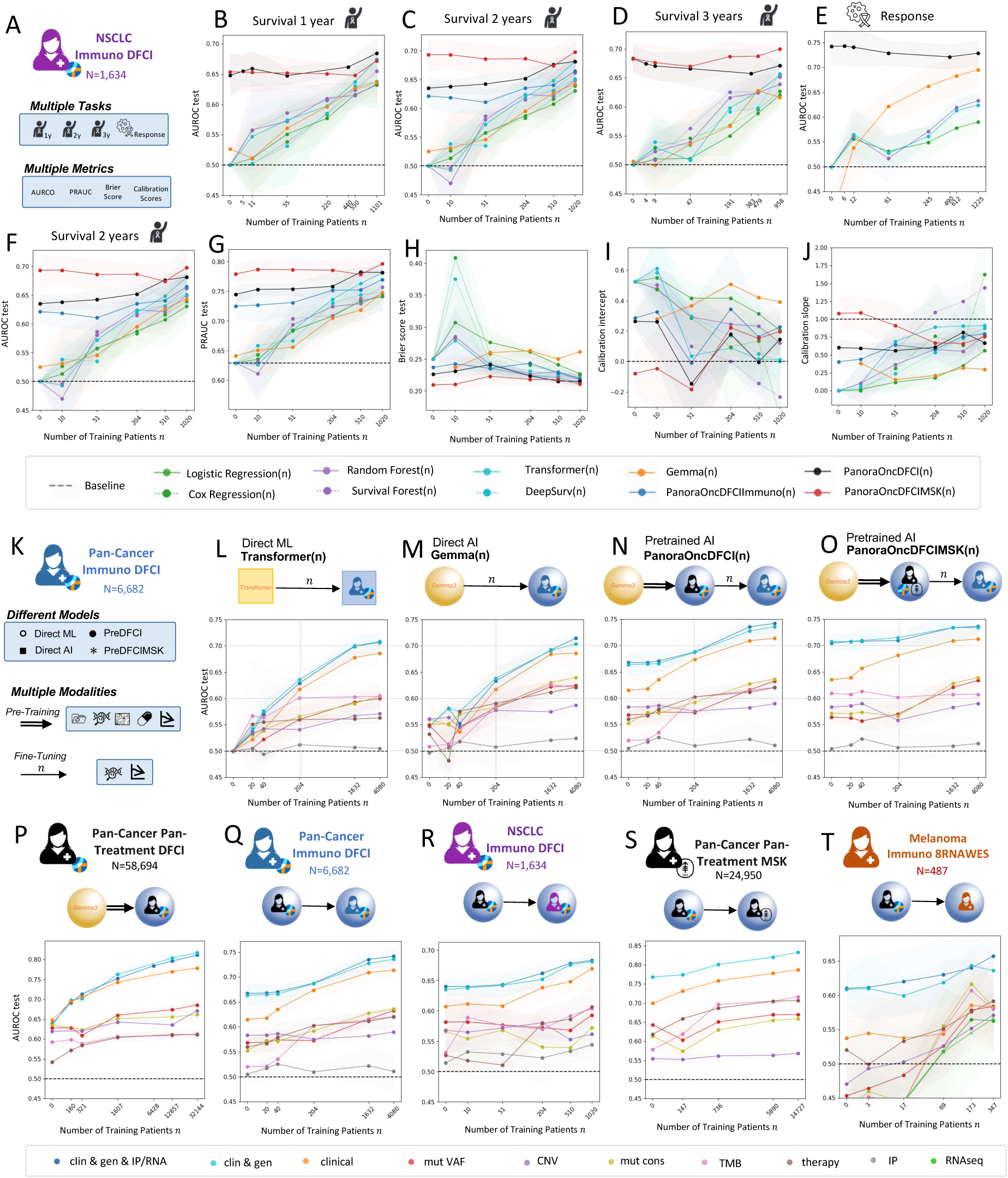
Performance analysis of PanoraOnc across multiple tasks, metrics, modalities, and cohorts. **(A)** Comparison of PanoraOnc performance on the DFCI NSCLC immuno cohort across different treatment outcome prediction tasks and evaluation metrics. **(B–E)** AUROC performance for one-, two-, and three-year survival prediction, as well as treatment response prediction, with increasing numbers n of training samples compared to different baseline methods. **(F–J)** Performance for two-year survival prediction evaluated using AUROC, PRAUC, Brier score, and calibration intercept and slope. **(K)** Analysis of the effects of direct training versus multimodal pretraining and modality importance for two-year survival prediction and the DFCI pan-cancer immune cohort. Comparisons include **(L)** a transformer-based machine learning model trained from scratch for each modality, **(M)** direct AI fine-tuning of Gemma(n) on individual modalities, **(N)** fine-tuning of individual modalities using the pretrained PanoraOncDFCI(n) model, and **(O)** fine-tuning using the pretrained PanoraOncDFCIMSK(n) model. AUROC performance of PanoraOncDFCI(n) on the internal **(P–R)** and external **(S–T)** cohorts across increasing numbers of training samples and different modality combinations is shown. Extended results for all outcomes, metrics, and cohorts, including comparisons with the survival, machine learning and direct AI fine-tuning, are presented in Figures S7, S8 and S9.

To quantify modality-specific contributions to predictive performance of the *PanoraOnc* models, we pretrained the models on the DFCI and MSK cohorts using all modalities and fine-tuned them using each modality separately or some combinations thereof (Fig. 3K), enabling the assessment of the relative importance of clinical, genomic, transcriptomic, and imaging data across cohorts and training set sizes. Specifically, we compared four training approaches using the DFCI pan-cancer immuno cohort: a machine learning model (Transformer^33^) trained from scratch for each modality (Fig. 3L), direct AI fine-tuning of individual modalities without pretraining, *Gemma(n)* (Fig. 3M), and fine-tuning of individual modalities using the pretrained *PanoraOncDFCI(n)* (Fig. 3N) and *PanoraOncDFCIMSK(n)* (Fig. 3O) models. In the zero-shot and few-shot regimes, pretrained models achieved substantially higher prediction accuracies across all or most modalities and converged to superior performance as the volume of training data increased (Fig. 3L-O). These results highlight the promise of large-scale pretrained AI models for robust multimodal learning from real-world patient data.

We further evaluated *PanoraOncDFCI(n)* when fine-tuned on internal (Fig. 3P–R) and external cohorts (Fig.3S-T). Although clinical and demographic features consistently contributed most to the predictive performance in both the zero-shot and asymptotic settings in the DFCI and MSK cohorts (Fig. 3P-T), incorporating genomic, transcriptomic, and imaging features in the multimodal model consistently improved overall performance. For instance, in the largest patient cohort (Fig. 3P), the overall AUROC reaches 0.82, constituting a +0.05 AUROC over clinical features alone 0.77. Nevertheless, the genomic modalities alone are predictive for survival, for instance, in the DFCI pan-cancer immune cohort (Fig. 3Q), the best genomic modality (mutation consequences) reaches an AUROC of 0.64 out of a total 0.74 for survival prediction. In the NSCLC cohort (Fig. 3R), the IP modality alone contributes up to 0.05 AUROC points and adds a small performance gain (0.02 AUROC points) over clinical and genomic features alone, suggesting that it provides some orthogonal information. When externally validating these results in the MSK cohort (Fig. 3S), we obtained robust zero-shot and few-shot performance among all modalities. When validating the model in the melanoma 8RNAWES cohorts (Fig. 3T), we found that the contribution of clinical features is smaller than in the DFCI cohorts, likely due to the reduced availability of clinical data in the 8RNAWES cohorts. Meanwhile, WES and RNA features alone approach the performance of clinical information, highlighting their information content and suggesting that pre-training on larger cohorts with WES and RNAseq datasets will further improve performance. Tumor mutational burden also remains a strong predictor in all cohorts, while treatment assignment effects revealed biases inherent to clinical trial data, as for instance observed in the 8RNAWES cohorts (Fig. 3T). These findings (Fig. 3, Fig. S8/S9) demonstrate that pre-training on real-world cancer cohorts with multiple modalities leads to improved performance gains in both zero-shot and asymptotic settings even when only data from single modalities are available.

### PanoraOnc identifies canonical and under-appreciated features associated with immunotherapy response

We next aimed to quantify the contributions of individual input features for outcome predictions to aid in patient stratification for clinical trials and biomarker discovery. To this end, we implemented an explainable AI framework for *PanoraOnc* using Integrated Gradients^34^ (IG)–based attribution methods, in which individual feature tokens are replaced with padding tokens (Fig. 4A). Feature contributions are then computed by integrating gradients from the padding embedding to the original input embedding with respect to the log-odds of the predicted survival label (Methods; Fig. S10). IG attribution scores were computed per feature and per patient and subsequently aggregated using the median to derive cohort-level feature importance rankings (Fig. 4B; Fig S10C,D; Methods). To incorporate broad molecular information for each gene, we aggregated the contributions from copy number variation (CNV), mutation consequence, mutation variant allele frequences (VAFs), and transcript abundance by RNA-seq. To validate these rankings and assess their dependence on both outcomes and feature values, we analyzed patient outcomes (Fig. 4C) and model-predicted survival probability distributions (Fig. 4D) across feature categories from internal and external cohorts.

**Figure 4:**
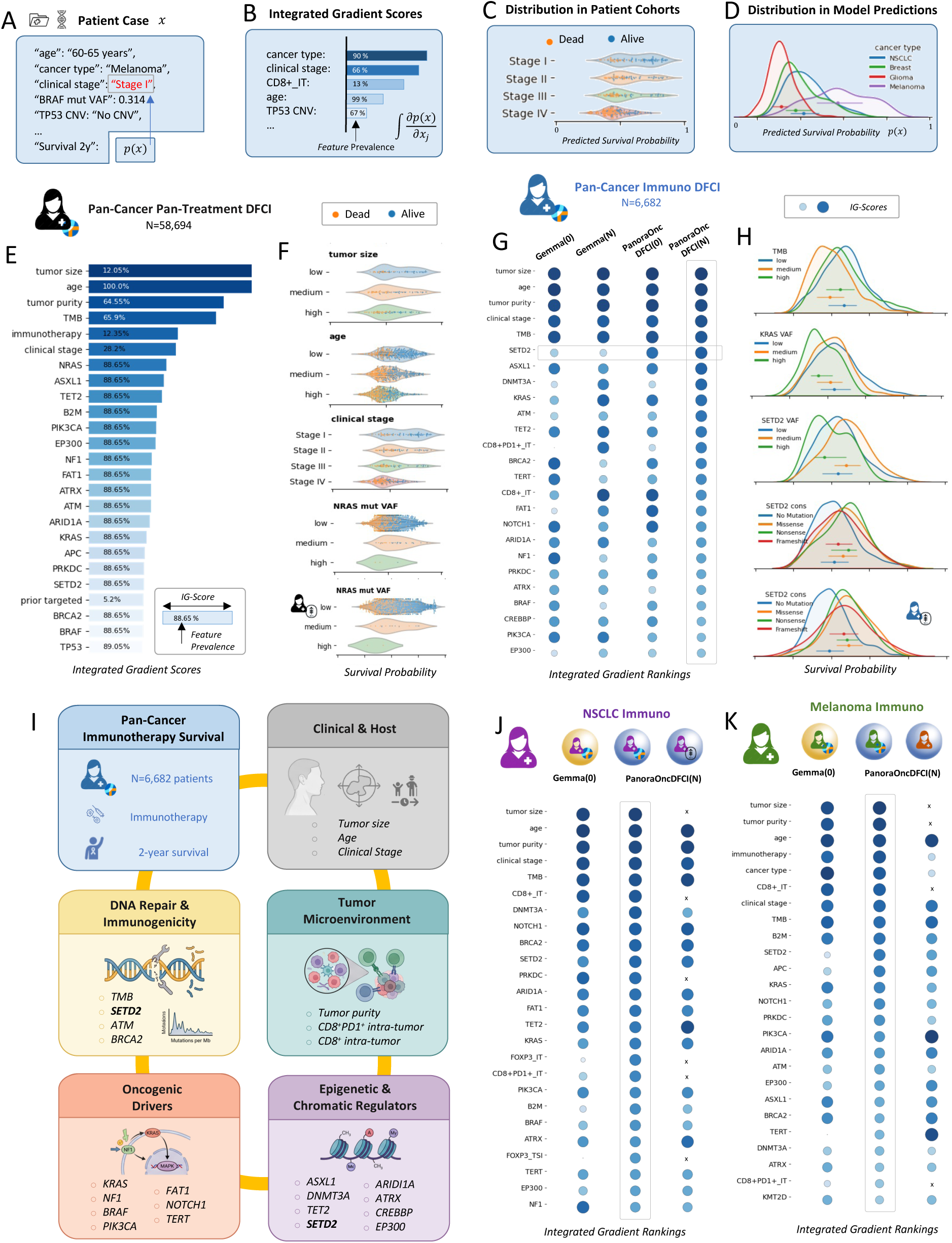
Quantitative Feature Attribution and Biomarker Discovery with PanoraOnc. **(A–D)** Overview of the Integrated Gradients (IG) framework for quantitative feature importance assessment in the PanoraOnc model. **(A)** demonstrates padding of individual features for calculation of **(B)** Integrated Gradient (IG) scores. These scores are used to rank features important for outcome predictions, which are subsequently investigated using the real-world data distributions **(C)** as well as **(D)** model-predicted survival distributions. **(E)** Top IG-ranked features for 2-year survival in the pan-cancer, pan-treatment DFCI cohort using the pretrained and fine-tuned PanoraOncDFCI(N) model. **(F)** Density plots and data distribution in the underlying data for selected features. **(G)** Evolving feature attributions for Gemma(0), Gemma(N), PanoraOncDFCI(0), and PanoraOncDFCI(N) models in the DFCI pan-cancer immunotherapy cohort. **(H)** Density plots highlighting well-known and underappreciated features for pan-cancer immunotherapy response. **(I)** Multi-axial categorization of PanoraOncDFCI(N)-identified features associated with pan-cancer immunotherapy survival, including clinical host factors, DNA repair and immunogenicity, the tumor microenvironment, oncogenic drivers, and epigenetic and chromatin regulators. **(J)** Feature importance rankings for the DFCI NSCLC cohort and **(K)** the melanoma immunotherapy cohort. Additional rankings for all cohorts are provided in Fig. S10.

Using this explainability framework, we systematically identified clinico-genomic features most influential for predicting outcomes following immunotherapy. Analyses focused on IG-ranked features for 2-year survival across cohorts (Figs. S10E-L). When evaluating the pretrained *PanoraOncDFCI(N)* model on the full DFCI cohort, top-ranked features included established clinical and biological factors such as tumor size, patient age, tumor purity, clinical stage, treatment type, and alterations or aberrant expression of cancer-associated genes including NRAS, PIK3CA, KRAS, APC, and TP53 (Fig. 4E). These features showed consistent patterns in both internal and external validation cohorts (Fig. 4F). We next examined how feature attributions evolved across model variants during pretraining and fine-tuning within the pan-cancer Immuno DFCI cohort (Fig. 4G) by comparing their aggregated IG scores at different training steps. Across *Gemma(0), Gemma(N), PanoraOncDFCI(0),* and *PanoraOncDFCI(N),* the highest-ranked features were consistent, reflecting shared learning of dominant clinical and biological signals. Notably, certain genomic features, such as SETD2, emerged prominently only in the pan-cancer–pretrained and fine-tuned models, but not in the untrained base model or in models trained solely on cohort-specific data. This observation suggests that pan-cancer pretraining facilitates learning of sparse, potentially nonlinear, and multimodal interactions across genomic modalities that are difficult to identify from limited cohort-specific data alone. To further explore these interactions, we analyzed raw data distributions and model predictions for top-ranked features in the pan-cancer immunotherapy cohort (Fig. 4H). Pretrained and fine-tuned models highlighted several well-established predictors of immunotherapy response, such as tumor mutational burden (TMB) and intratumoral CD8⁺ T-cell infiltration, as well as multiple factors without a well-established role in immunotherapy response, including SETD2, DNMT3A, FAT1, and EP300. These features were weakly ranked or absent in the general-purpose base model but consistently ranked higher in the pan-cancer–pretrained PanoraOnc models, suggesting that pretraining on pan-cancer and pan-treatment cohorts improves sensitivity to such signals.

We then evaluated IG rankings from *PanoraOncDFCI(N)* within individual cancer-specific cohorts. In the NSCLC cohort (Fig. 4I–J), alongside canonical factors such as clinical stage, TMB, B2M, and CD8⁺ T-cell infiltration, the model highlighted multiple novel features related to signal transduction (NOTCH1, KRAS), tumor-infiltrating clonal hematopoiesis (DNMT3A), and epigenetic remodeling (ATRX, EP300). In the melanoma immunotherapy cohort (Fig. 4K–L), prominent features included treatment type, CD8⁺ T-cell infiltration, TMB, B2M, and KRAS followed by features with a potentially underappreciated role in immunotherapy response, such as ARID1A, APC, ATM, EP300, and TERT. Across pan-cancer immunotherapy cohorts, influential features spanned multiple biological axes, including clinical burden (tumor size, stage), host characteristics (age), tumor composition (purity, immune cell infiltration), and tumor-intrinsic genomic programs. Measures related to immunogenicity and DNA damage response (e.g., TMB, ATM, BRCA2, SETD2) ranked highly, alongside epigenetic regulators and genes frequently associated with clonal hematopoiesis (ASXL1, DNMT3A, SETD2). Canonical oncogenic drivers such as KRAS and TERT also contributed, consistent with their known immune-modulatory roles^35–38^. Overall, these results support the view that immunotherapy response and survival are shaped by integrated clinical, immune, and genomic factors rather than single biomarkers.

### PanoraOnc enables identification of patient subcohorts that may benefit from alternative treatments

Beyond patient stratification and biomarker discovery, *PanoraOnc* can be used to identify patient subgroups that may benefit from alternative therapeutic strategies. Specifically, our framework estimates differential treatment outcomes across treatment categories by predicting both factual outcomes (under the treatment actually received) and the counterfactual treatment outcomes (under hypothetical alternative treatments). These hypothetical treatments can correspond to broad therapeutic categories, such as chemotherapy, immunotherapy, or targeted therapy, or can be extended to individual therapeutic agents. To infer hypothetical treatment outcomes from real-world observational data, we developed a bias-adjusted counterfactual framework based on a doubly-robust approach^39,40^ (Methods). We applied this framework to the quantitative two-year survival predictions generated by the pretrained *PanoraOncDFCIMSK(N)* model for all NSCLC patients in the DFCI and MSK cohorts (N = 6,282) who received chemotherapy, immunotherapy, or targeted therapy (Fig. 5A). We first examined predicted survival probabilities under the factual treatment assignment policy, i.e. predictions for each patient under the treatment they received in the observational dataset (Fig. 5B), yielding factual survival distributions (Fig. S11F). Because observational data are subject to treatment assignment biases (Methods, Fig. S11B, S11J, S12C), we next generated counterfactual outcome predictions from *PanoraOncDFCIMSK(N)* for each patient under all alternative treatment categories (Fig. 5C). These predictions were then adjusted using *targeted maximum likelihood* (TMLE^39,40^, Methods), a doubly robust method from causal treatment effect estimation that combines the model-based outcome predictions with information from the observed treatment assignment probabilities to produce bias-aware counterfactual treatment outcome estimates (Fig. S11L). Intuitively, TMLE “corrects” the predicted outcomes by accounting for how likely each patient was to receive each treatment in the real-world data, ensuring that survival estimates reflect both the predictive model and the actual treatment assignment patterns. Based on these bias-adjusted counterfactual outcomes, we identified the top-ranked treatment category for each patient (Fig. 5C), enabling stratification of the NSCLC cohort into factual subgroups and identification of patients who would potentially have achieved better outcomes under alternative treatments (Fig. 5D). Applying this approach to the NSCLC cohort revealed several subgroups of interest (Fig. 5E), including patients whose factual treatment was ranked highest by the model (diagonal panels) and patient groups who may warrant investigation of alternative therapies (off-diagonal panels). We then analyzed significantly different features for each factual–counterfactual pair to characterize patient subgroups potentially benefitting from alternative treatments (Figs. 5F–I, Fig. S12D). A subgroup of patients treated with chemotherapy may benefit from switching to targeted therapies if they have later stage tumors with EGFR mutations, while patients with KRAS mutations, primary tumors, and higher TMB may fare better remaining on chemotherapy (Fig. 5F). Similarly, a subgroup of patients receiving immunotherapy may achieve better outcomes with chemotherapy if older or if the biopsy sample was obtained from the primary tumor, while later-stage patients with KRAS or TP53 mutations and male sex may benefit from remaining on immunotherapy (Fig. 5G). For targeted therapy, patients with primary tumor biopsies and high tumor purity might benefit from switching to chemotherapy (Fig. 5H), while others may switch to targeted therapy based on EGFR mutations, primary tumor biopsies, and older age, whereas patients with KRAS, NF1, FAT1, KMT2D, TP53 mutations, and larger TMB are predicted to benefit from remaining on immunotherapy (Fig. 5I). To further characterize these subgroups, we analyzed distributions of individual therapy agents (Fig. S13A), prior treatments (Fig. S13B), clinical stage (Fig. S14A), number of prior treatment regimens (Fig. S14B), different biopsy sites (Fig. S15A), and different specific mutations profiles such as EGFR (Fig. S15B), highlighting nuanced differential capabilities. Overall, these analyses demonstrate the ability of *PanoraOnc* to identify patient subgroups who may benefit from treatment categories different from those they actually received (the factual treatment), such as chemotherapy, immunotherapy or targeted therapy. Together, these results demonstrate that transferable predictions derived from large-scale multimodal pretraining can enable bias-aware exploration of treatment outcomes, providing a foundation for *in silico* clinical trials with interpretable predictions under real-world data heterogeneity and scarcity in precision oncology.

**Figure 5:**
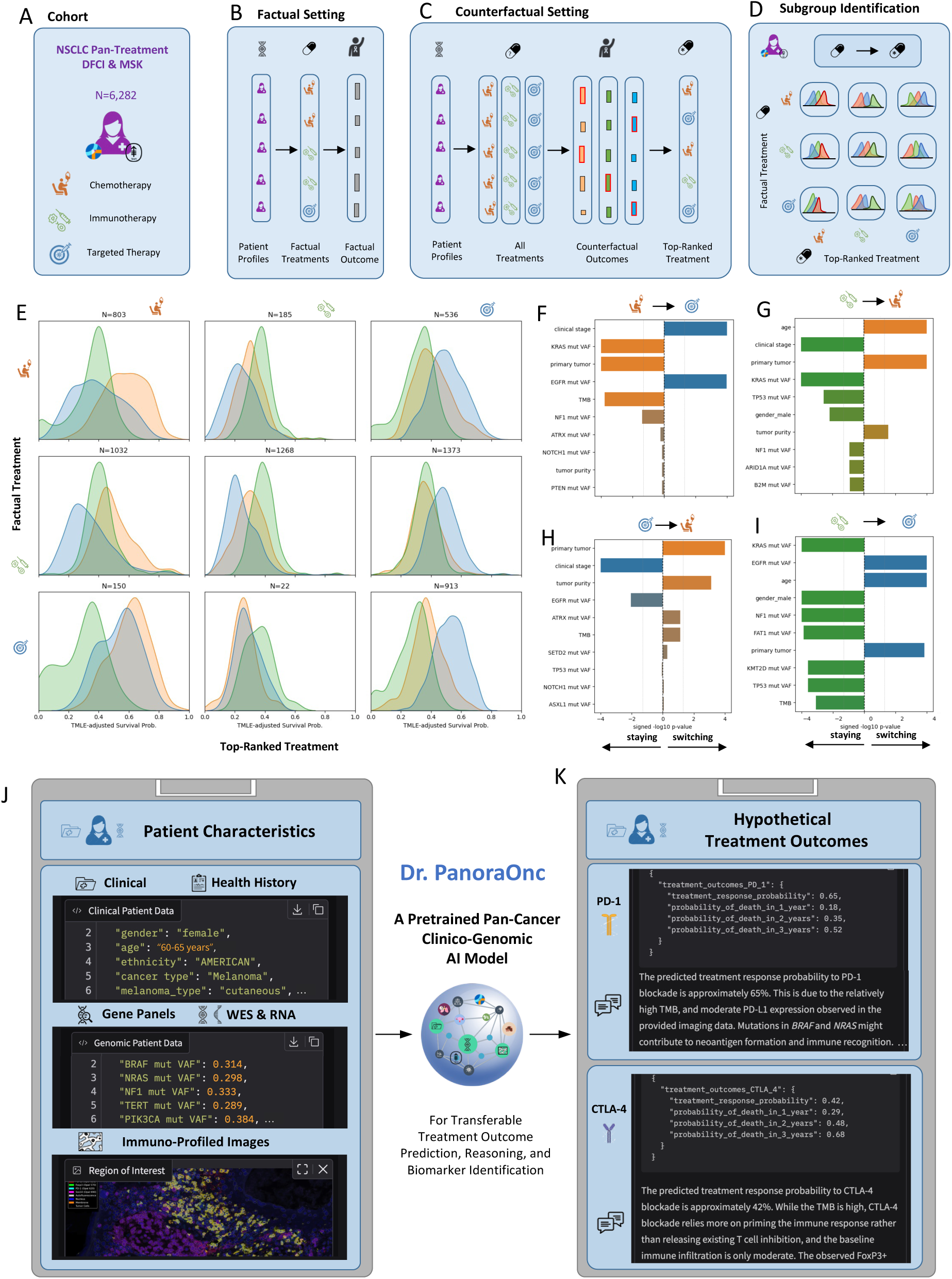
Bias-Aware Counterfactual Analysis and Interactive Agent Dr. PanoraOnc. **(A)** Counterfactual treatment analysis across patients from the pan-treatment NSCLC cohorts from MSK and DFCI, treated with either chemotherapy, immunotherapy, and targeted therapy. **(B)** For each patient, factual treatment outcome survival probabilities were estimated using PanoraOncDFCIMSK and compared to **(C)** counterfactually propensity-adjusted, doubly robust counterfactual outcome estimates, enabling bias-aware ranking of hypothetical treatment scenarios (Methods, Figs. S11/S12). **(D)** This framework allows stratification of the pan-treatment NSCLC cohort and identification of subgroups that may have achieved better outcomes under alternative treatments, highlighting candidates who might benefit from treatment switching. **(E)** Stratified subpopulation survival probability distributions, where rows represent the factual treatments received and columns indicate the top-ranked treatment category. The diagonal represents patients for whom the factual treatment remains optimal, while off-diagonal entries indicate patients who may potentially benefit from an alternative treatment. **(F–I)** For each factual–counterfactual subcohort, key driving features were analyzed to characterize patient subgroups likely to benefit from alternative treatments. More details over the subcohorts are provided in Figures S13-S15. **(J)** Dr. PanoraOnc, an interactive clinico-genomic AI agent built on PanoraOncDFCIMSK, enabling scenario-based treatment outcome prediction and qualitative reasoning for real-world patient cases. **(K)** Example application to a synthetic melanoma patient integrating clinical, genomic, and immune imaging data, producing quantitative and qualitative counterfactual treatment predictions. The complete example is provided in Fig. S16.

Finally, building on our pretrained *PanoraOnc* model, we developed a prototypical interactive AI agent, *Dr. PanoraOnc* (Fig. 5J-5K, Fig. S16), that generates hypotheses about personalized patient outcomes from multimodal clinico-genomic data. The agent enables both quantitative and qualitative exploration of predicted treatment outcomes, including the investigation of counterfactual therapeutic scenarios for individual cancer patients.

## Discussion

Our study demonstrates the potential of a pretrained clinico-genomic AI model to advance personalized oncology by integrating multiple, heterogeneous clinico-genomic cancer patient cohorts at scale and transferring learned knowledge to new cohorts from other hospitals or smaller, clinically focused cohorts. By pretraining on large real-world pan-cancer clinico-genomic datasets encompassing 66 cancer types, we showed that a vision language model–based approach achieves significant zero- and few-shot performance compared to state-of-the-art methods, including external zero-shot validation for quantitative treatment outcome predictions, allowing transfer of the pretrained information to other hospitals, cohorts, or therapeutic settings. Moreover, our approach can integrate many different clinico-genomic multimodal data modalities into a unifying AI framework for multi-task treatment outcome prediction and is readily extensible to additional modalities not included in the present study, such as proteomics. A central strength of our approach lies in its ability to unify different heterogenous cohorts with many features and modalities into a single transferable predictive framework and enabling the identification of both established and novel biomarker candidates of immunotherapy response and survival in the context of the high-dimensional multimodal patient characteristics. Our pretrained AI framework is especially valuable for few-shot learning settings such as for rare cancer types, knowledge- and data-scarce settings such as early-phase clinical trials with limited sample sizes, offering a principled framework for sample acquisition, in-context biomarker discovery, and cohort stratification. Finally, the demonstrated zero- and few-shot transferability across cancer types and institutions may be particularly impactful for resource-limited hospitals, including those in low- and middle-income countries, which often lack the infrastructure to build large local clinico-genomic databases and could benefit substantially from transferable pretrained models to advance personalized oncology.

Despite these advances, several limitations point to important avenues for future work. First, although we leveraged two of the largest available pan-cancer clinico-genomic cohorts, generalizability to completely new patients from different hospitals is constrained by the breadth and representativeness of the available datasets; additional large-and small-scale cohorts for pre-training, fine-tuning and cross-evaluation, including those from diverse hospitals from different countries and underrepresented subgroups, will be essential to fully realize AI models that generalize well in new settings. Second, while broad pan-cancer pretraining substantially improved few-shot performance, asymptotic gains were moderate, motivating exploration of enhanced pre-training and transfer learning strategies such as contrastive multimodal learning and domain-adaptive losses^41^. Third, our AI framework primarily leverages the semi-structured demographic, clinical, treatment, imaging, genomic, and transcriptomic data, with candidate features selected through knowledge- and data-driven approaches. Towards more comprehensive multimodal medical AI systems, future work will extend beyond the use of the most informative features per modality and incorporating pretrained modality-specific backbone models for fully exploiting the high-dimensional input space. For instance, including pretrained backbone models in the VLM for RNA-seq^42^, and additional modalities such as unstructured reports^9^, histopathology^15^, radiology^13^, single-cell transcriptomics^43^, spatial transcriptomics^44^ and proteomics^45,46^, as well as ensembles of AI agent approaches^47,48^ could offer additional opportunities for richer patient representations, deeper mechanistic insights, and improved performance accuracy.

Prognostic zero-shot generalization, as demonstrated in this study, represents an important first step toward transferable and clinically actionable outcome prediction across hospitals, cancer types, and treatment settings. However, extending such predictive generalization toward treatment-specific, patient-level recommendations remains challenging due to systematic differences in clinical guidelines, historical standards of care, and treatment assignment practices across institutions with several treatment assignment biases^4,49^ and additional challenges due to unmeasured confounding^4^. Addressing these challenges in future work through advanced counterfactual and causal inference techniques^4,50^ such as other doubly-robust propensity corrections^51^, difference-in-differences^52^ and synthetic controls^52^, together with uncertainty quantification methods^53,54^ will be essential to improve robustness, interpretability, and clinical reliability. Ultimately, such advances directly included in pretrained AI models could enable the simulation of in-silico virtual clinical trials, which will require rigorous retrospective, prospective, and interventional validation before clinical deployment.

Moreover, due to privacy, ethical, and regulatory constraints, pretrained AI models containing sensitive clinical and genomic patient data cannot be directly shared openly. Approaches leveraging cross-pretrained federated learning^55^ protocols with model-sharing agreements, together with the integration of differential privacy into AI foundational models^56^, represent promising directions to build shareable multimodal AI foundation models. Several other practical considerations must be addressed to translate these methods into clinical impact in the future. Although the models’ quantitative predictions were thoroughly benchmarked, robust evaluation of the performance of the interactive AI agent Dr. PanoraOnc (Fig. S16) and its reasoning processes remains an important direction for future work to ensure interpretability and trust. IG-based feature analyses revealed potential biomarker candidates requiring prospective validation in more independent cohorts. Finally, endpoint heterogeneity across datasets, for instance, RECIST-based response definitions versus NLP-annotated response labels^9^, or differences in survival reference points introduces label noise that limits predictive accuracy and interpretability. Expanding prediction targets to adverse events and competing risks, combined with longitudinal^57^ and interventional patient trajectories modelling, will be crucial towards building a comprehensive AI-powered virtual patient.

Deployable AI systems for personalized oncology critically depend on the ability of models to generalize reliably to new clinical settings, including unseen hospitals, cancer types, and treatment regimens, without direct access to matched patient cohorts. The continued development of pretrained pan-cancer AI foundation models with transferable treatment outcome prediction capacities, such as *PanoraOnc*, points toward a future in which personalized oncology is powered by AI systems pretrained on large, international cohorts that integrate clinical, genomic, and imaging data directly into clinical workflows and clinical trial design. Such models can further incorporate complementary sources of clinical and biomedical knowledge, including predefined therapy rules derived from medical guidance, curated clinical trial datasets, and mechanistic insights from patient-derived ex-vivo or in vitro perturbation data such as cell lines, xenografts, and organoids. Integrating these heterogenous data sources enables the learning of comprehensive treatment–response representations within a pretrained unified AI framework. This paradigm facilitates the development of end-to-end in-silico lab-clinic loop AI systems that connect pre-clinical experiments, molecular profiling, medical imaging, and real-world patient data. When further refined through active and reinforcement learning^58^, such in-silico AI systems could provide the foundation for AI-driven digital medical twins that support individualized oncology decision-making, inform optimized treatment strategies, and advance precision medicine at both the molecular and patient levels in the future.

## Methods

### Data alignment, harmonization, and preprocessing

We curated multimodal clinical–molecular datasets for the DFCI^17–19,59^ (Figs. S1-S4, [Tables S1-S4](https://drive.google.com/drive/folders/1f_Nikfb3fj5WohAAfw4kfm-x0guVYKnc?usp=sharing)), MSK^20,60^ (Fig. S5, [Tables S5-S7](https://drive.google.com/drive/folders/1f_Nikfb3fj5WohAAfw4kfm-x0guVYKnc?usp=sharing)) and 8RNAWES^21–28,61^ (Fig. S6, [Table S8](https://drive.google.com/drive/folders/1f_Nikfb3fj5WohAAfw4kfm-x0guVYKnc?usp=sharing)) cohorts, comprising electronic health records (demographics, clinical variables, treatments, outcomes), targeted sequencing panels and whole-exome sequencing (WES) including somatic mutation consequences (MUT), variant allele frequencies (VAFs), and copy number alterations (CNV), together with RNA sequencing (RNA-seq) and immunohistochemistry-profiled images (IP) images. Each patient data is abstractly represented as a triplet (**x**, **a**, **y**) ∈ X × A × Y, where we define an anchoring time point *t_0_* corresponding to a clinically relevant treatment decision (Fig. 2B). Information prior to *t_0_* forms the high-dimensional and multimodal patient profile

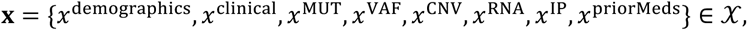

whereas information after *t_0_* constitutes the treatment outcome profile **y** ∈ Y. The treatment options available at *t_0_* are represented by **a** ∈ A. All clinical, genomic, and transcriptomic features in the DFCI, MSK, and 8RNAWES cohorts were standardized and preprocessed using harmonized pipelines to ensure consistency and comparability for subsequent ML and AI modeling.

### Clinical and demographic data

The DFCI, MSK, and 8RNAWES cohorts included structured clinical and demographic features extracted from electronic health records (institutional databases at DFCI and MSK) and study metadata (8RNAWES). Patient demographic features *x^demo^*^g*rap*ℎ*ics*^ comprised age, gender, ethnicity, education level, and marital status and lifestyle factors such as tobacco and alcohol use. Clinical features *x^clinical^* captured cancer type, disease stage, biopsy site and type, and tumor size. Across DFCI, MSK and 8RNAWES cohorts, feature names and their categories were standardized and harmonized to enable consistency across cohorts.

### Genomic data

Genomic features for solid tumors were derived from high-coverage targeted sequencing panels: DFCI OncoPanel^18^ (v1: 275 genes, 200x coverage; v2: 302 genes, 350x; v3: 447 genes, 350x) and MSK IMPACT^62^ (341-505, 750x coverage). For hematological cancer types, genomic features were obtained from specialized blood panels: DFCI RapidHeme v2 (95 genes, 1500x) and v3 (88 genes, 325x), and MSK ImpactHeme^63^ (400 genes, 750x coverage). Whole-exome sequencing (WES) data in the 8RNAWES cohort were harmonized to the DFCI gene panel using the OncoPanel pipeline^18^. For each gene, DNA-derived features included somatic mutations using functional consequences *x*^MUT^ and variant allele frequency *x*^VAF^, together with copy number variations *x*^CNV^ and tumor-level summary metrics such as tumor mutational burden (TMB) and tumor purity. Mutation categories were harmonized across DFCI, MSK, 8RNAWES cohorts following OncoPanel definitions^18^, with “No Mutation” assigned to wild-type genes. In cases of multiple mutations per gene, the mutation with the highest VAF was retained. CNVs were also categorized across cohorts per standard OncoPanel conventions: 0 = *Two-copy or Homozygous Deletion*, 1 = *One-copy or Heterozygous Deletion*, 2 = *No CNV* 3–5 = *Low-level Gain*, and ≥6 = *High-level Amplification*, with 8RNAWES CNVs preprocessed using FACETS v3.6.3^64^. Due to the overabundance of genomic features, the analysis focused on the 100 most frequently altered 100 genes for each genomic modality (mutation consequences, mutation VAFs, and CNVs).

### Transcriptomic data

RNA sequencing (RNA-seq) data were obtained from the Genotypes and Phenotypes (dbGaP) sequencing repository or the European Genome-Phenome Archive (EGA) repositories for all 8RNAWES^21–28^ cohorts and processed using the RNA-seq Immune Analysis (RIMA) pipeline^65^. This included alignment to GRCh38 reference genome using STAR^66^ v2.6.1 and transcript quantification in transcripts per million (TPM) using Salmon^67^ v0.13.1. Gene-level TPM values were directly used as transcriptomics features *x*^RNA^ and compared to knowledge-driven derived features *x*^RNApre^ such as normalized expression signatures for inflammation (CD274, CD8A, LAG3, STAT1), tertiary lymphoid structures (TLS; CD79B, CD1D, CCR6, LAT, SKAP1, CETP, EIF1AY, RBP5, PTGDS), and antigen presentation (B2M, HLA-A, HLA-B, HLA-C, HLA-E, HLA-F, HLA-G, HLA-H, TAP2). Immune cell composition was estimated using CIBERSORTX^68^ (LM22 reference), quantifying 22 immune cell types, and T-cell and B-cell repertoires were analyzed using TRUST4^69^. Additional features included TIDE-derived scores^70,71^ reflecting T-cell dysfunction, immune exclusion, IFNG signaling, and immune checkpoint activity (CD274, CD8, MDSC, CAF, TAM.M2, CTL). Together, these transcriptomic features provide both raw expression profiles and biologically informed signatures of the immune landscape and tumor microenvironment, harmonized across patients for integrative multimodal modeling.

### Multiplex immunofluorescence imaging

We aligned the clinical and genomic patient data with spatially and single-cell resolved multiplex immunofluorescence (IF) imaging data. Whole-slide images include DAPI, FOXP3, CD8, PD-1, PD-L1, and autofluorescence channels, together with a tumor-specific marker: cytokeratin for epithelial cancers, PAX8 for renal cell carcinoma, and SOX10 for melanoma. The whole-slide images were cropped into three to six local regions of interest^19^ per patients. Derived features *x*^IP^ capture intra-tumor (IT), tumor–stromal interface (TSI), and combined metrics, e.g., CD8+_IT, PD1+_TSI, FOXP3+_IT, PD-L1_IT/TSI expression across regions^19^.

### Treatments

Treatment information **a** ∈ A was aligned to a treatment decision point *t_0_* (Fig. 2B). For patients treated with immunotherapy, the anchoring time *t_0_* corresponded to the first immune checkpoint therapy administered in the patient trajectory, stratified into PD-1 (nivolumab, pembrolizumab, cemiplimab), CTLA-4 (ipilimumab, tremelimumab), and PD-L1 (atezolizumab, durvalumab, avelumab) including combinations. For non-immunotherapy-treated patients, the anchoring time *t_0_* corresponds to the first therapy initiated post-genomic profiling, categorized as chemotherapy, targeted therapy, biological therapy, hormonal therapy, or bone-marrow modifying agents, following established categorization of treatment agents^20^. The timepoint of genomic profiling was represented by the date of “sample ordering” in the DFCI cohorts, while it was “study initiation” in the 8RNAWES cohorts, and “genomic profile reporting” in the MSK cohorts. Prior treatment medication categories *x*^priorMeds^ relative to anchoring time *t_0_* were included in the patient profile *x*. Any features such genomic or imaging information collected after treatment initiation were excluded. Temporal differences between genomic biopsy and treatment initiation were encoded as additional features into the patient profile.

### Treatment Outcomes

Treatment outcomes are defined as **y** = (*y^D^*^1^, *y^D^*^2^, *y^D^*^3^, *y^R^*) ∈ Y, where *y^Dt^* = **1**(*D* ≤ *t*) represents cumulative mortality within *t* ∈ {1,2,3} years after the anchoring time point *t_0_*, and *D* is a random variable representing time to death. Specifically, we predict a binary outcomes for a fixed-horizon *t*, where 1 denotes patient death within *t* years, and 0 denotes documented survival beyond *t* years. Patients with unknown survival status (e.g. lost to follow-up or censored due to study cutoff) before year *t* were treated as censored and encoded as missing for the corresponding outcome *y^Dt^*, and were excluded from the training for that outcome. Survival labels were obtained from original study metadata for 8RNAWES cohorts. For the DFCI and MSK cohorts, death events and observed death times were derived from the overall survival and last alive data. The treatment response variable *y^R^* ∈ {0,1} represents binary response status, corresponding to binarized RECIST^72^ (v1.1) response labels for responders (partial or complete response) versus non-responders (stable or progressive disease) in the 8RNAWES cohorts, and NLP-derived labels^9^ for the DFCI cohort, while missing for the MSK cohorts.

### Patient representation and treatment outcome prediction

Each patient is modeled as a triplet of random variables (**X**, **A**, **Y**) ∈ X × A × Y. The objective is to estimate the multivariate conditional probability distribution *μ**_a_***_,***y***_(**x**) = ℙ[***Y*** = ***y*** ∣ **X** = **x**, ***A*** = **a**] for treatment outcomes ***y*** ∈ Y given patient characteristics **x** ∈ X and treatment options **a** ∈ A. Given multimodal patient data 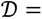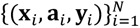 for patient *i* in a cohort with *N* samples, these probabilities are learned from observed real-world factual patient cases *μ**_a_******_i_***_,***y***_***_i_***(**x***_i_*) = ℙ[***Y*** = ***y_i_*** ∣ **X** = **x***_i_*, ***A*** = **a_i_**] using statistical, ML, or AI methods. This framework enables comparison of hypothetical treatment outcomes *μ**_a_*****_1_**_,***y***_(**x***_i_*), *μ**_a_*****_2_**_,***y***_(**x***_i_*), … across different treatment options ***a***_1_, ***a***_2_, …for a patient profile **x***_i_*.

### AI model training

Our *PanoraOnc* models are built upon the open-weight vision–language model Gemma3-12B^29^, initialized with 12 billion pretrained parameters Θ_0_. The model is based on a decoder-only transformer architecture and adopts a multimodal design with a transformer-based text encoder and a SigLIP^73^ visual backbone, which itself is a Vision Transformer^74^, pretrained using a CLIP-style^75^ contrastive loss to align visual and textual embeddings in a shared latent space. This pretrained architecture enables joint reasoning over text and image modalities. For *PanoraOnc* models, further pretraining and task-specific fine-tuning were performed on multimodal clinical, genomic, and imaging-derived patient data using Low-Rank Adaptation (LoRA^76^) with trainable parameters *θ*_0_. LoRA enables parameter-efficient adaptation while mitigating catastrophic forgetting of previously acquired knowledge. To reduce memory footprint and training costs, we employed a 4-bit quantized implementation from *unsloth*^77^ of Gemma3-12B. All experiments were implemented in *PyTorch*^78^. During both pretraining and fine-tuning, the visual backbone remained frozen (since we train on image-derived features, not raw images) and only LoRA adapters in the language model component were updated. All patient input modalities (clinical, genomics, transcriptomics, images, treatment, outcomes) were tokenized and embedded into the shared transformer latent space, enabling unified multimodal representation learning across cohort datasets.

### Patient case representation for the AI model

Each patient case (**x**, **a**, **y**) ∈ X × A × Y was encoded in a JSON-like format *json*(**x**, **a**, **y**), capturing quantitative and qualitative information while preserving contextualized feature names (Fig. S10A). This representation leverages prior knowledge embedded in the pretrained *Gemma3* model, including medical literature and ontologies, and reduces inconsistencies in naming and missingness patterns across cohorts. This JSON representation was tokenized using the *SentencePiece*^72^ tokenizer from Gemma3-12B^29^, yielding a sequence of tokens **s** = **s**_1:J_ = R*s*_1_, …, *s_J_*S ∈ ℤ*^J^*, where each token s_j_ ∈ {0,1, …, |V| − 1} for a vocabulary V and maximum token length *J* ranged from 2,048 to 4,096 tokens depending on the number of modalities and features in the cohort. The tokenized sequence **s** of each patient is then processed by the pretrained vision–language encoder to produce hidden representations **s** ↦ **z** = **z_1_**_:**J**_ = R**z_1_**, …, **z_J_**S ∈ ℝ*^J^*^×*E*^ with token embeddings **z_j_** ∈ ℝ*^E^* of fixed embedding dimension *E* = 3840 as specified in Gemma3. These embeddings integrate structured, semi-structured, and image-derived information into a unified representation that can be continuously pretrained and fine-tuned with real-world patient cohorts. Instruction fine-tuning was performed using a chat-template^29^ format combining multimodal inputs with a system prompt and task specification (e.g., treatment response or survival at one, two, or three years), enabling the model to generate quantitative and qualitative treatment outcomes directly in a textual and interactive form without an additional classification head (Fig. S10B).

### Loss functions and training objective

Training followed a standard autoregressive next-token prediction objective. At each position *j*, the hidden state **z**_j_ ∈ ℝ*^E^* was projected into vocabulary logits **η**_j_ ∈ ℝ^|V|^, where |V| denotes the vocabulary size. The conditional probability *p_θ_*(*s*_j+1_ = *v* ∣ **s**_1:j_) = *p_θ_*(*s*_j+1_ = *v* ∣ **z**_1:j_) of the next token *s*_j+1_ equals any *v* ∈ V was then obtained via *softmax* transformation, *p_θ_*(*s*_j+1_ = *v* ∣ **s**_1:j_) = *softmax*(**η**_j_)[*v*]. During pretraining, the cross-entropy loss was applied across all tokens in the patient sequence, 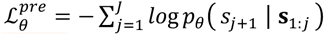 yielding pretrained weights 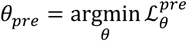, initialized from *θ*_0_. For supervised fine-tuning on scalar patient outcomes *y_i_*, corresponding to the binary treatment outcomes, the loss was applied only at the target token position *k* + 1, that is, 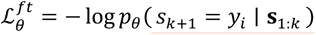, conditioned on the tokenized patient profile and treatment (**x***_i_*, **a***_i_*) ↦ **s**_1:*k*_ ∈ ℤ*^k^*. For direct fine-tuning, the stochastic optimization problem 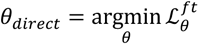 was solved with initialization with *θ*_0_, whereas for fine-tuning after pre-training, the weights were initialized with *θ_pre_*. All optimization problems were solved using stochastic gradient descent (ADAM^79^) with mini-batch updates over multiple training epochs, with gradients computed via backpropagation and parameters updated using adapted learning rate (see details below). This unified generative formulation enables calibrated quantitative prediction directly in the PanoraOnc model *p_θ_*, that is, we calibrated the probabilities *p_θ_*(*s_k_*_+1_ = *y_i_* ∣ **s**_1:*k*_) = ℙ[*Y* = *y_i_* ∣ **X** = **x***_i_*, ***A*** = ***a****_i_*] based on many real-world patient cases 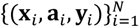, while maintaining compatibility with qualitative and interactive language outputs. In contrast, traditional machine learning baselines were trained on tabular representations of (**x***_i_*, **a***_i_*) derived separately from each modality together with scalar labels *y_i_*, offering limited generalization between tasks, datasets and outcomes.

### AI model training, and evaluation

For each cohort D_c_of size *N*_c_ (for instance *c* = DFCI Immuno cohort with *N_c_* = 6,682), we performed four-fold cross-validation. In each fold, 25% of the patients (0.25 *N*_c_) were held out as a fixed test set 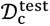, while the remaining 75% (0.75 *N*_c_) formed the training pool 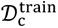. Each quarter served once as test set. This repeated splitting reduces variance in performance estimates, particularly in cohorts with limited sample size or imbalanced outcomes. From each training pool, models were trained on varying fractions *n* of available training data, ranging from zero-shot (*n* = 0) to the full training set (*n* = 0.75 *N*_c_). All evaluations were performed on the same fixed test set within each fold. The base model *Gemma3* was evaluated directly on 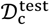 without task-specific training. For direct fine-tuning, *Gemma(n)* was trained on n samples from 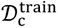 and evaluated on 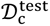. To prevent data leakage between pretraining and fine-tuning stages, complementary exclusion cohorts were constructed via set subtraction, that is, D_DFCI_No_ *_c_* = D_DFCI_ ∖ D_c_. For example, D_DFCI_NoImmuno_ = D_DFCI_ ∖ D_DFCI_Immuno_ with analogous constructions for the other sub cohorts. Combined datasets were defined via set union, e.g., D_DFCI_MSK_ = D_DFCI_ ∪ D_MSK_ for the joint DFCI and MSK cohorts. All pretraining and fine-tuning experiments explicitly excluded overlapping patients between pretraining and fine-tuning cohorts. For pretraining-transfer experiments such as *PanoraOncDFCI(0)*, the model was first pretrained on an exclusion cohort e.g. 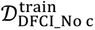, and then evaluated on the corresponding fixed test set 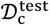. For the pretrained and fine-tuned experiments such as *PanoraOncDFCI*(n), the model was first pretrained on an exclusion cohort 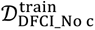, then fine-tuned on n samples from 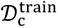, and evaluated on the corresponding fixed test set 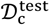. For example, when fine-tuning on the DFCI NSCLC immunotherapy-treated cohort D_DFCINSCLC Immuno_, all NSCLC immunotherapy patients were excluded from the pretraining stage to prevent overlap, such that D_DFCI_NoNSCLC Immuno_ = D_DFCI_ ∖ D_DFCI_NSCLC Immuno_ was used for pretraining in this experiment. Identical train–test splits were used for both direct and transfer fine-tuning to ensure comparability. For each cohort, split, modality configuration, and training fraction, models were trained independently from scratch (except when initialized from pretrained parameters). Consequently, thousands of *PanoraOnc* models were trained to generate the results presented in Figs. 2 and 3. This two-phase training design enables cross-cancer generalization while preserving sensitivity to specific clinical tasks.

### Hyperparameter and GPU setup

Pretraining and fine-tuning were performed with per-device batch sizes of 16 and 8, respectively, combined with gradient accumulation steps of 2 and 4 to fit within the memory constraints of NVIDIA A100 (80 GB) and NVIDIA H100 (94 GB VRAM) GPUs. All experiments were conducted on internal GPU servers equipped with 4× NVIDIA H100 and 4× NVIDIA A100 (80 GB) accelerators. Training, inference, generation, and evaluations were performed using 4-bit quantization via the *Unsloth*^77^ framework to reduce memory footprint and computational cost. The maximum token length *J* per patient case ranged from 2048 to 4096 tokens, depending on cohort modality richness. Models were trained for 4–12 epochs in pretraining and direct fine-tuning experiments, as specified per experiment in the accompanying code. For transfer learning experiments involving pretraining followed by task-specific fine-tuning, fine-tuning was performed only for 2 epochs in each cohort. The learning rate was set to 2 × 10^−4^ with weight decay 0.01, and 5 warmup steps were used for the learning rate scheduler. LoRA^76^-based fine-tuning employed configurations *r* = 8, *α* = 8 and *r* = 16, *α* = 16 for task-specific adaptation. Additional ablation studies evaluated the impact of varying learning rates, weight decay, and LoRA parameters. Base models of different sizes (4B, 12B, and 27B parameters) were also assessed. Although minor differences were observed in the zero-shot setting, continued training resulted in no statistically significant performance differences across model sizes. For consistency, all reported experiments were conducted using the 12B base model.

### Statistical, survival, and machine learning baselines

We benchmarked *PanoraOnc* models against a range of classical statistical, machine learning, and survival models implemented using the *scikit-learn*^80^, *PyTorch*^78^, and sklearn-survival^81^. Baseline statistical models included logistic regression and regularized logistic regression^82^ (Lasso and Ridge) to capture linear relationships and promote sparsity. To model nonlinear feature interactions, we evaluated ensemble and neural approaches, including Random Forest^83^, Light Gradient Boosting Machine (LGBM^80^) and multilayer perceptron (MLP^80^). We further compared against TabPFN^33^, a pretrained transformer-based tabular foundation model designed for robust performance across structured datasets. For survival models, we used a linear Cox proportional hazard model^30^, as well as the nonlinear models Random Survival Forest^31^ and DeepSurv^32^. These approaches represent strong baselines for quantitative outcome prediction in the survival literature and machine learning literature, particularly in high-dimensional feature spaces^84^. Continuous variables were standardized to zero mean and unit variance, and categorical variables were one-hot encoded with a designated reference category removed to avoid perfect collinearity. Missing values were handled using cohort-specific imputation strategies consistent across all baseline models (median for continues features, and additional *None* category for categorical features).

### Metrics for quantitative evaluation of probabilistic forecasts

For classical statistical and machine learning models, predicted probabilities were directly used to evaluate binary classification discrimination performance. We computed the area under the receiver operating characteristic curve (AUROC) and the precision-recall area under the curve (PRAUC) to assess discriminative ability to emphasize performance under class imbalance, particularly for low-prevalence outcomes. To further evaluate survival and calibrations validity, we computed Brier scores at different survival points (one- to three-year-survival), as well as calibration curves represented via fitted calibration slopes (optimal is 1) and intercepts (optimal is 0). For the VLM-based AI models, treatment outcomes were encoded as textual tokens *v* ∈ {*True*, *False*} at predefined target positions in the output sequence. During evaluation, we extracted the logits at the designated label position *k+1*, that is *p_θ_*(*s_k_*_+1_ = *True* ∣ **s**_1:*k*_) and *p_θ_*(*s_k_*_+1_ = *False* ∣ ***s***_1:*k*_). For binary fine-tuning tasks, probabilities were normalized 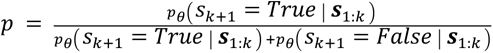 to obtain calibrated class probabilities. These probabilities were then used to compute the different evaluation metrics, enabling direct quantitative comparison between the generative language model and conventional statistical, machine learning, and survival approaches.

### Interpretability method for VLM models

To quantify feature importance in our *PanoraOnc* models, we implemented Integrated Gradients (IG^34^) from the *Captum*^85^ package and wrapped it around the pretrained *PanoraOnc* models. IGs are a path-integrated gradient attribution method that mitigates gradient saturation effects observed at convergence. Let a patient case (**x***_i_*, **a***_i_*, y_i_) be represented by hidden embeddings **z** = **z_1_**_:**k**_ = R**z**_1_, …, ***z***_j_, …, **z**_k_S ∈ ℝ*^k^*^×*E*^corresponding to the tokenized patient case (**x***_i_*, **a***_i_*) ↦ ***s***_1:*k*_ = R*s*_1_, …, *s*_j_, …, *s_k_*S ∈ ℤ*^k^*, where ***z***_j_ ∈ ℝ*^E^* denotes the embedding of token *s*_j_. Let 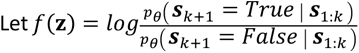 denote the trained scalar prediction function at position *k* + 1 corresponding to the log-odds of the predicted binary outcome y_i_ (e.g., survival at 2 year) obtained from a trained *PanoraOnc* model *p_θ_*. We define the baseline embedding **z**^′^ ∈ ℝ*^k^*^×*E*^ comprising all the padding-tokens. For each input token *s*_j_, the Integrated Gradients attribution is defined as 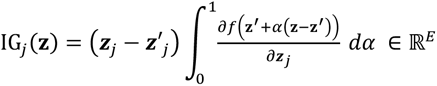. This formulation computes the path integral of the gradient along the straight-line interpolation from the baseline embedding **z**^′^ to the original embedding **z**. The resulting attribution vector IG_j_(**z**) ∈ ℝ*^E^* quantifies the contribution of token *s*_j_ to the model’s prediction at token *s_k_*_+1_. For scalar feature importance per input token *j*, we computed the median absolute attribution across embedding dimensions, 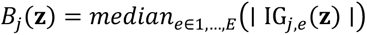, which provides robustness to extreme gradient values. These attribution scores were computed for all patient indices *i* = 1, . . ., *N* and input token indices *j* = 1, . . ., *k*. Because multiple tokens may correspond to a single clinical or molecular feature (e.g., gene name, mutation annotation, RNA value), we aggregated token-level attributions to feature-level scores. For a feature *g* represented by token index set J_g_, the feature attribution for patient *i* is *B_i_*_,g_ = Σ_j∈J_*_g_B*_j_ (**z***_i_*). For gene-level summaries, contributions from copy-number variation (CNV), mutation consequence, mutation VAF, and RNA-seq tokens corresponding to the same gene were further aggregated by 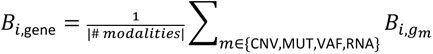 Cohort-level feature importance rankings were obtained by aggregating patient-level attributions using the median 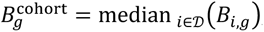, which provides robustness to outliers and skewed attribution distributions and were used for ranking the features.

### Bias-aware doubly robust counterfactual adjustment

Assume a real-world observational patient cohort 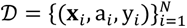 with N patients comprising patient profiles ***x****_i_* ∈ X, observed factual treatments *a_i_* ∈ {*a*_1_, …, *a_C_*} = A and observed real-world outcomes y_i_ ∈ {0,1} (e.g. y = 1 represents 2-year survival). We denote the outcome predicted by *PanoraOnc* as *μ_a_*_,*y*_(***x****_i_*) = E[Y ∣ *X* = ***x****_i_*, *A* = *a*] = ℙ[Y = 1 ∣ *X* = ***x****_i_*, *A* = *a*], which estimates the expected outcome under treatment *a* ∈ A for a patient with covariates ***x****_i_*. The treatment-specific average prediction under the factual treatment policy, restricted to patients who actually received treatment *a*, is 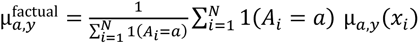. This estimator (Fig. S11E) reflects the empirical treatment allocation distribution (Fig. S11F) in the cohort and is biased in the presence of non-random treatment assignment. A model-based counterfactual estimate instead averages predicted hypothetical outcomes over the full cohort 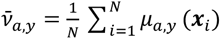 (Fig. S11G). This approach removes explicit conditioning on the observed treatment and estimates the mean of the hypothetical outcome under treatment *a* according to the outcome model. When evaluating *PanoraOncDFCI(N)* in this counterfactual setting, by predicting outcomes *μ_a_*_1,*y*_(**x***_i_*), *μ_a_*_2,*y*_(**x***_i_*), … for each patient **x***_i_* ∈ D under all alternative treatment categories *a* ∈ A, the resulting survival distributions for a fixed outcome *y* (e.g. 2-year survival) can be determined (Fig. S11H). However, these individualized counterfactuals remain sensitive to misspecification of *μ_a_*_,*y*_(*x*) since they do not explicitly adjust for treatment assignment bias. Treatment assignment bias^4,49^ reflects conditionally imbalanced treatment probability distributions, where ℙ(*A* = *a_k_* ∣ *X* = ***x****_i_*) ≠ ℙk *A* = *a*_g_ ∣ *X* = ***x****_i_* l for any *a_k_*, *a*_g_ ∈ A, *k* ≠ *g* and for any ***x****_i_* ∈ X. Consequently, such individualized counterfactual predictions remain model-based and are not identifiable without additional assumptions. To first characterize treatment assignment biases, we estimated the underlying (average) treatment assignment policy and estimated treatment propensities *e_a_*(***x****_i_*) = ℙ(*A* = *a* ∣ *X* = ***x****_i_*) (Fig. S11I) using multivariate logistic regression trained on clinical and genomic covariates, including demographics, disease stage, cancer type, prior therapies, tumor mutational burden, and recurrent oncogenic alterations. The resulting propensity distributions (Fig. S11J) reveal overlap between the treatment categories, and the subsequent analyses were conducted on a trimmed cohort 0.01 < *e_a_*(*x_i_*) < 0.99, to remove extreme cases^4^. To obtain bias-adjusted population-level counterfactual outcomes, we applied a targeted maximum likelihood estimation (TMLE^39,40,86^), which is doubly robust estimator^51,87^ (Fig. S11K) TMLE starts from the treatment outcome model *μ_a_*_,*y*_(**x**) obtained from *PanoraOnc* and performs a targeted update using information from the propensity model *e_a_*(***x***) =. Specifically, for each treatment *a*, we define the clever covariate 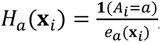, and fit a fluctuation (targeting) model that updates the initial predictions via a logistic regression, that is, logit(*μ̃_a_*_,*y*_(**x***_i_*)) = logit(*μ_a_*_,*y*_(**x***_i_*)) + *ε* ⋅ *H_a_*(**x***_i_*), where *ε* is estimated from the data. This step explicitly corrects the initial outcome predictions using the observed outcomes and the propensity-weighted residuals^39,40,86^. The final TMLE estimate of the average potential outcome under treatment *a* is then given by the plug-in estimator 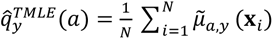. TMLE is doubly robust^51,87^, meaning it yields consistent estimates if either the outcome model *μ_a_*_,*y*_(**x**) or the propensity model *e_a_*(**x**) is correctly specified, assuming no unmeasured confounding and sufficient overlap (*e_a_*(**x***_i_*) > 0). In addition, TMLE ensures that predicted outcomes remain within valid bounds (e.g., probabilities between 0 and 1) and improving finite-sample stability. The resulting adjusted estimates provide bias-aware counterfactual outcome distributions (Fig. S11L). Comparing factual and top-ranked treatments (Fig. S11M) enables estimation of potential cohort level improvements (Fig. S11N). We use *PanoraOnc* to generate personalized hypothetical survival trajectories on the patient level, while TMLE adjusted treatment outcomes provides validated population- and sub-group level treatment effects (Fig. S11O) enabling identification of subcohorts that might benefit from alternative treatments (Fig. S11P). Future work will incorporate this causal adjustment directly into the pretrained AI model.

### Treatment subgroup identification and treatment recommendation analysis

This counterfactual framework (Figs. S11) allows stratification of the pan-treatment NSCLC cohort and identification of subgroups that may achieve better outcomes under alternative treatments, highlighting candidates who might benefit from treatment switching (Figs. S11P, S12A). In the resulting table, rows represent the factual treatments received, and columns indicate the top-ranked counterfactual treatment category. For each subcohorts, we estimate the potential improvement in outcomes under the top-ranked alternative treatments (Fig. S12B), and the factual propensity distribution (Fig. S12C), reflecting the current treatment policy. Moreover, for each factual–counterfactual subcohort, we characterize key driving features using a multivariate logistic regression (Fig. S12D). Let *c_i_* = 1 indicate that patient *i* should switch to the counterfactual treatment, and *c_i_* = 0 indicate staying on the factual treatment. The probability of switching is modeled as logit 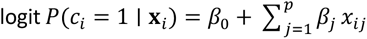 are the clinico-genomic features of patient *i*. Coefficients *β*_j_ were tested using t-statistics and corrected for multiple comparisons via the Benjamini–Hochberg false discovery rate (FDR) method. We then computed the signed of the negative logarithms of the adjusted p-values, that is *signed_score*_j_ = *sign*(*β*_j_) ⋅ −*log* _10_(*pval*_j,_*_adj_*), and plotted these values to rank the importance of each feature in each subcohort (Fig. S12B). In this context, the sign of *β*_j_ indicates the direction of association: a positive *β*_j_ implies that higher values (or presence) of the feature are associated with a higher probability of switching to the counterfactual treatment, whereas a negative *β*_j_ indicates association with staying on the factual treatment. To further characterize the different subgroups, we analyzed the distributions of the individual therapy agents at the decision point t_0_ (Fig. S13A), prior treatment categories (Fig. S13B), clinical stage (Fig. S14A), number of prior treatment lines (Fig. S14B), biopsy sites (Fig. S15A), and specific EGFR mutations (Fig. S15B). Prior lines of treatment were determined according to the medication administration dates in DFCI and MSK. Any gap of more than 30 days between administration of categorized agents was considered the start of a new line of treatment. This analysis highlights and characterizes patient subgroups that may benefit from alternative treatments.

### Interactive pan-cancer clinical-genomic AI agent Dr. PanoraOnc

To translate our pretrained and fine-tuned *PanoraOnc* model series into an interactive research platform, we developed *Dr. PanoraOnc*, an interactive multimodal AI tool for quantitative and qualitative treatment outcome prediction and reasoning. The tool, implemented using the *Gradio*^88^ framework, provides a conversational chat interface with the pretrained vision–language models *PanoraOncMSK(N) or PanoraOncDFCIMSK(N)*, accepting structured and free-text, clinical variables, genomic, transcriptomic, and imaging inputs (Fig. 5J-5K, Fig. S16). Users can input real-world patient cases, explore alternative treatment scenarios, and obtain individualized quantitative and qualitative predictions. This tool is intended for research use only and is not validated for clinical deployment, which will require retrospective, prospective, and interventional evaluation in the future^89^. *Dr. PanoraOncMSK* is an open-weight model, while *Dr. PanoraOncDFCIMSK* remains private-weight due to patient consent and IRB constraints.

## Data availability

Publicly available datasets include 5RNAWES^21–25,61^ and 3RNA-only^26–28^ melanoma cohorts, as well as MSK cohorts^20^, accessible via public repositories^60^. Clinical, demographic, and genomic data from DFCI, including OncoPanel sequencing^59^ and immune-profiled imaging (IP^19^) data, are available upon request from the corresponding authors. Access requires Institutional Review Board (IRB) approval for analysis of clinical, genomic, and imaging data from DFCI.

## Ethics/IRB

IRB approval was obtained for large-scale analysis of clinical and genomic features at DFCI, including treatment outcomes and NLP-derived response annotations as well as to integrate multiplex immunofluorescence imaging data with clinical and genomic data. Controlled-access datasets, such as 8RNAWES, were obtained according to repository-specific guidelines. Preprocessed and deidentified MSK clinical–genomic data are available without IRB requirements.

## Code availability

Code for data preprocessing, model pretraining and fine-tuning, experiments, benchmarking, integrated gradient analysis, and the interactive AI tool will be released upon publication via Github. Due to patient privacy, consent and data-use restrictions, pretrained and fine-tuned model weights based on DFCI patient data cannot be shared; only *PanoraOncMSK*, based on publicly available MSK data, has been released.

## Author contributions

MS, JG, CF, and FM conceived the study, with FM supervising all steps of the project. MS and JG performed data harmonization, alignment, cleaning, and preprocessing across all datasets. JG supported transcriptomic data preprocessing and tested the codebase. MS implemented all AI and ML models and conducted all experiments for PanoraOnc. CF provided advice on biological relevance and interpretation of findings and biomarkers. ABB provided baseline results for the survival models. TM contributed guidance on modeling and statistical interpretation. MS developed the interactive AI agent, Dr. PanoraOnc. JA provided expertise on image data access and integration. CG tested the codebase and contributed to improving the manuscript. JG, SM, GA, and GB supported genomic data harmonization and preprocessing. DL and KK advised on medical content and clinical relevance. MS generated figures under FM’s guidance and with input from all co-authors. MS and FM wrote the manuscript, with input and feedback from all co-authors. All authors contributed to interpreting results and refining the manuscript.

## Acknowledgements

We thank the Michor lab for helpful discussions and comments.

## Conflicts of interest

F. M. is a co-founder and advisor to Harbinger Health, an advisor to Zephyr AI, and a director of Recursion Pharmaceuticals. None of these relationships are directly or indirectly related to the contents of this manuscript. All other authors declare no conflicts.

---

**Figure S1:**
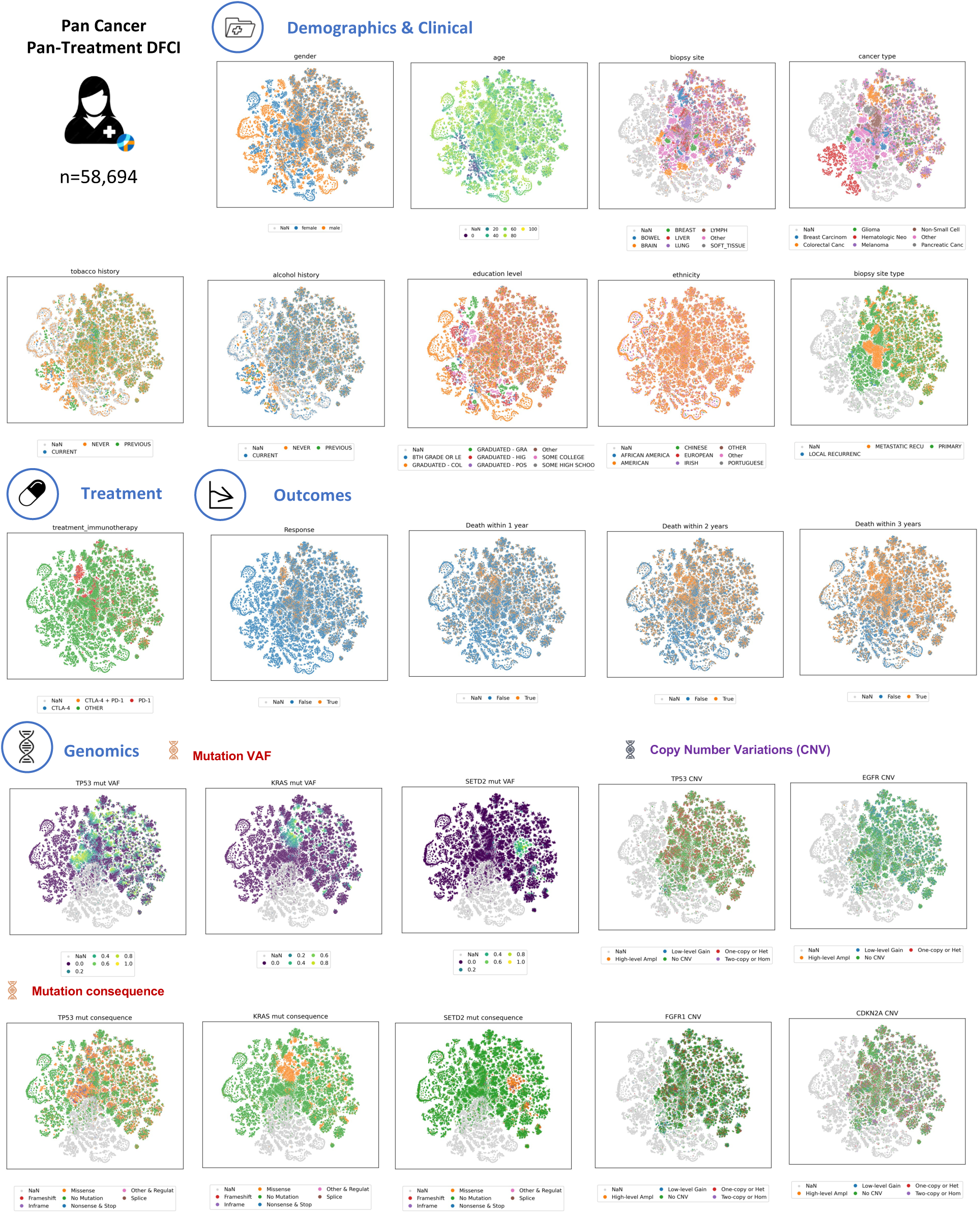
T-SNE Plots of the Pan-Cancer Pan-Treatment DFCI Cohort. Overview of all patients in the pan-cancer pan-treatment cohort in the t-SNE space, colored by a subset of selected features. Cohort summary is provided in Supplementary Table T1.

**Figure S2:**
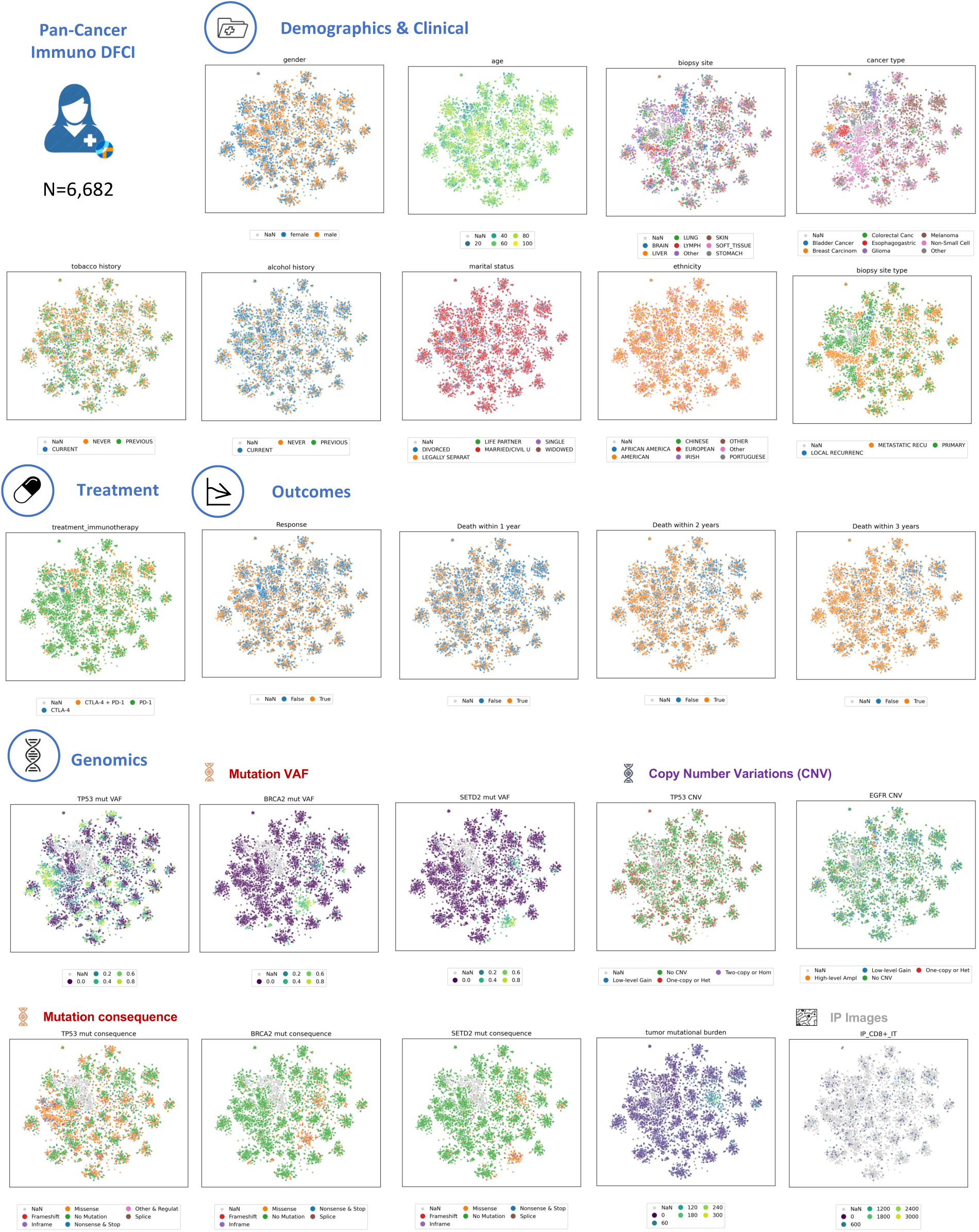
T-SNE Plots of the Pan-Cancer Immuno DFCI Cohort. Overview of all patients in the pan-cancer immuno treatment cohort in the t-SNE space, colored by a subset of selected features. Cohort summary is provided in Supplementary Table T2.

**Figure S3:**
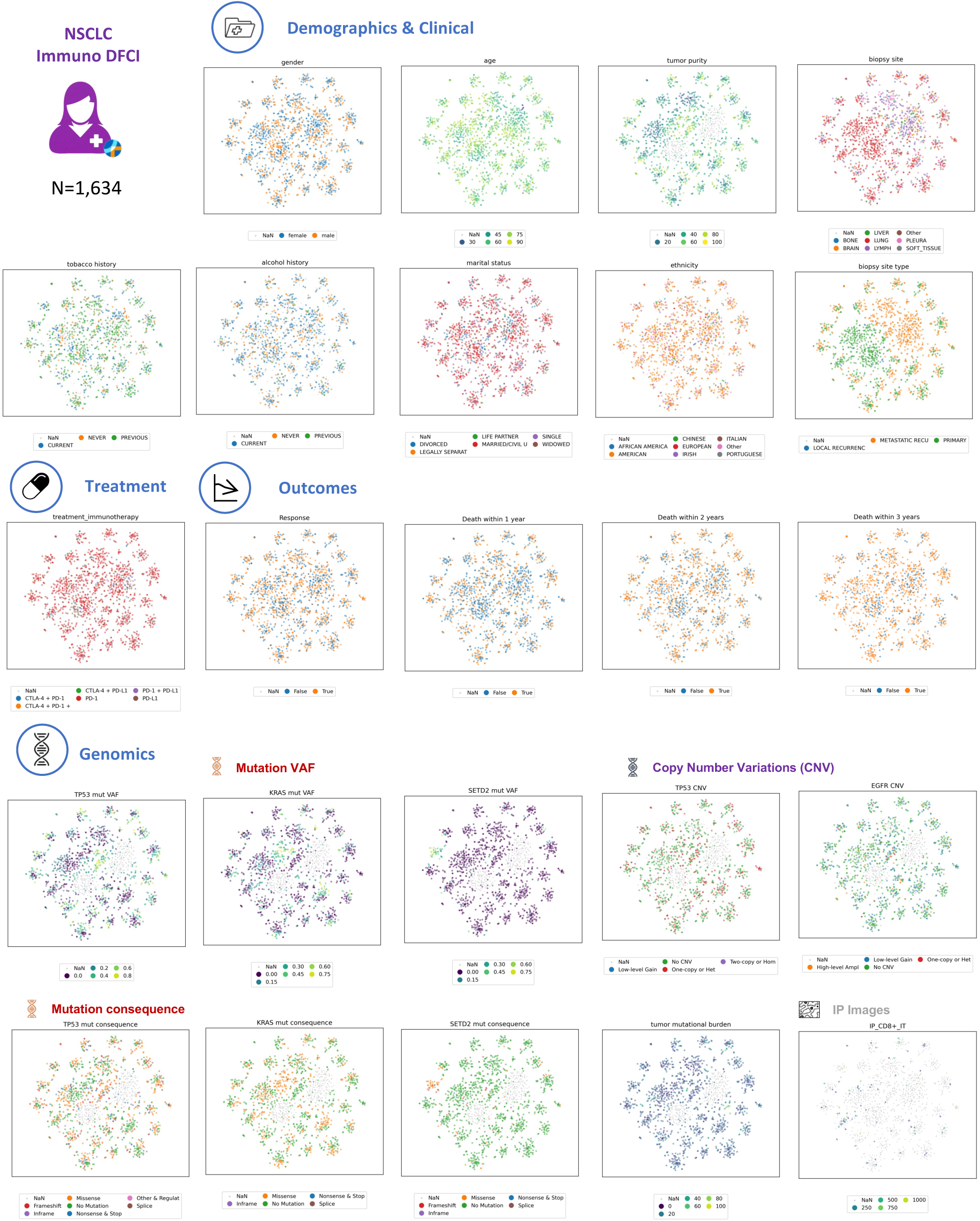
T-SNE Plots of the NSCLC Immuno DFCI Cohort. Overview of all patients in the non-small cell lung cancer (NSCLC) immunotherapy treatment cohort in the t-SNE space, colored by a subset of selected features. Cohort summary is provided in Supplementary Table T3.

**Figure S4:**
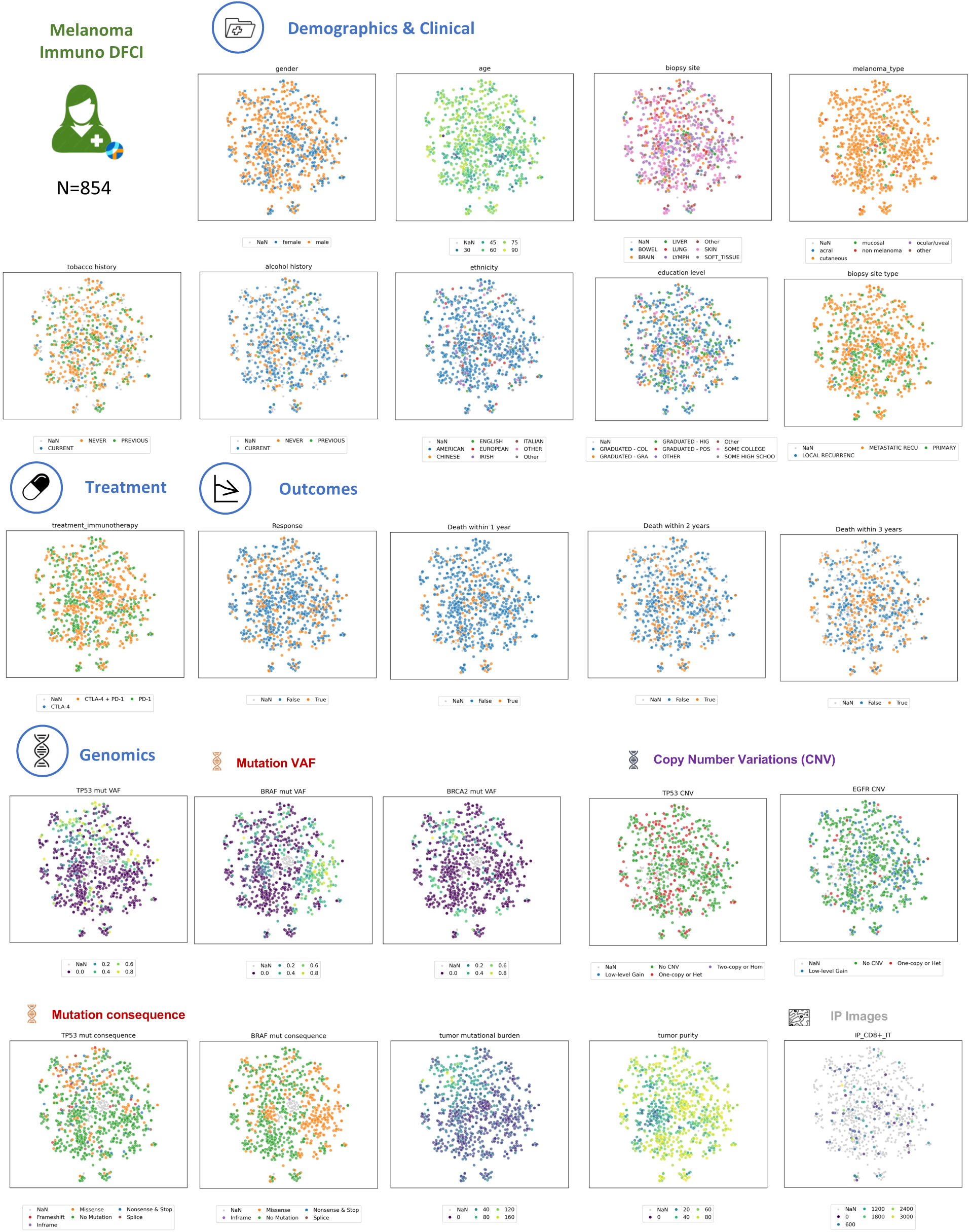
T-SNE Plots of the Melanoma Immuno DFCI Cohort. Overview of all patients in the melanoma immune treatment cohort in the t-SNE space, colored by a subset of selected features. Cohort summary is provided in Supplementary Table T4.

**Figure S5:**
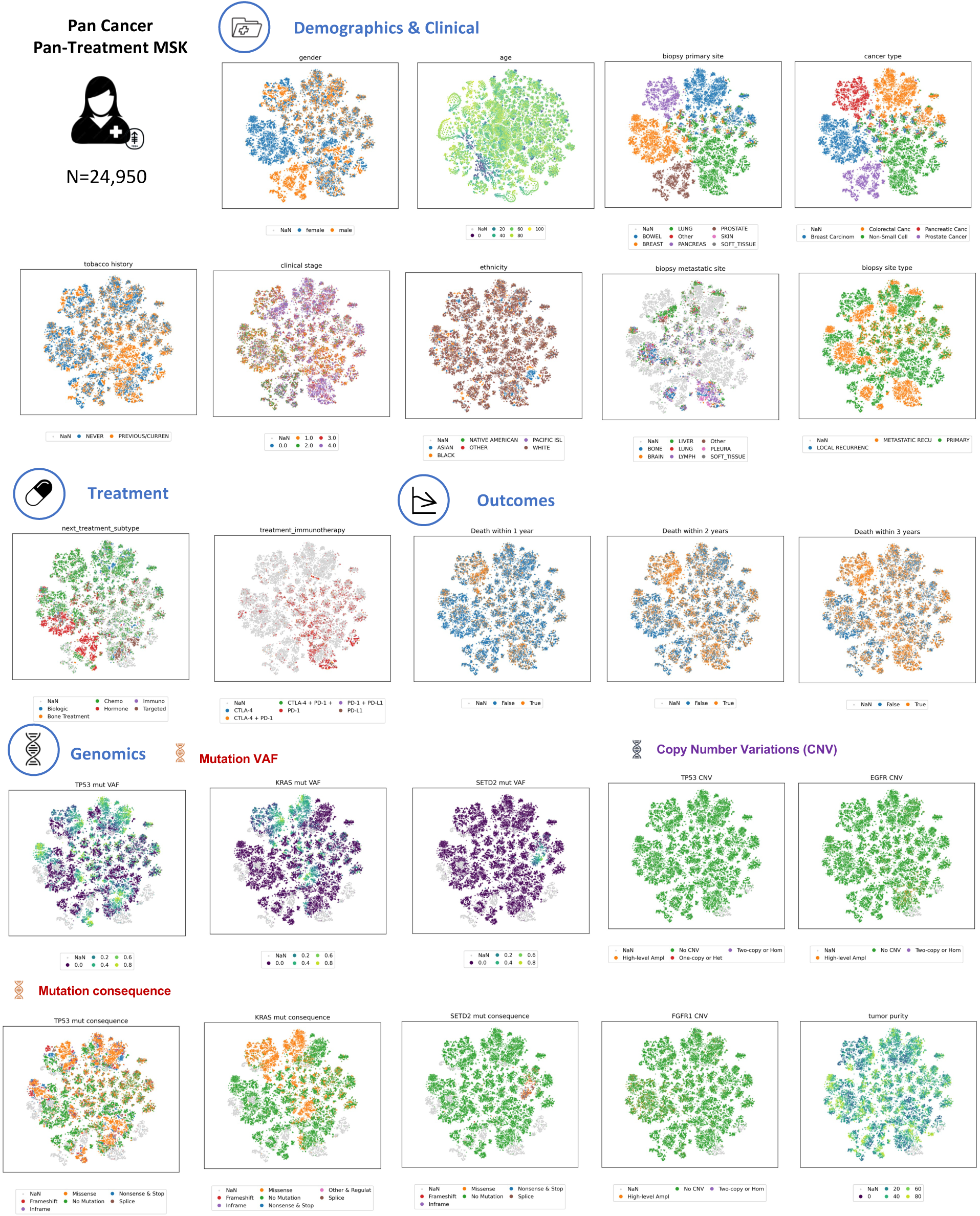
T-SNE Plots of the Pan-Cancer Pan-Treatment MSK Cohort. Overview of all patients in the pan-cancer pan-treatment MSK cohort in the t-SNE space, colored by a subset of selected features. Cohort summaries are provided in Supplementary Tables T5-T7.

**Figure S6:**
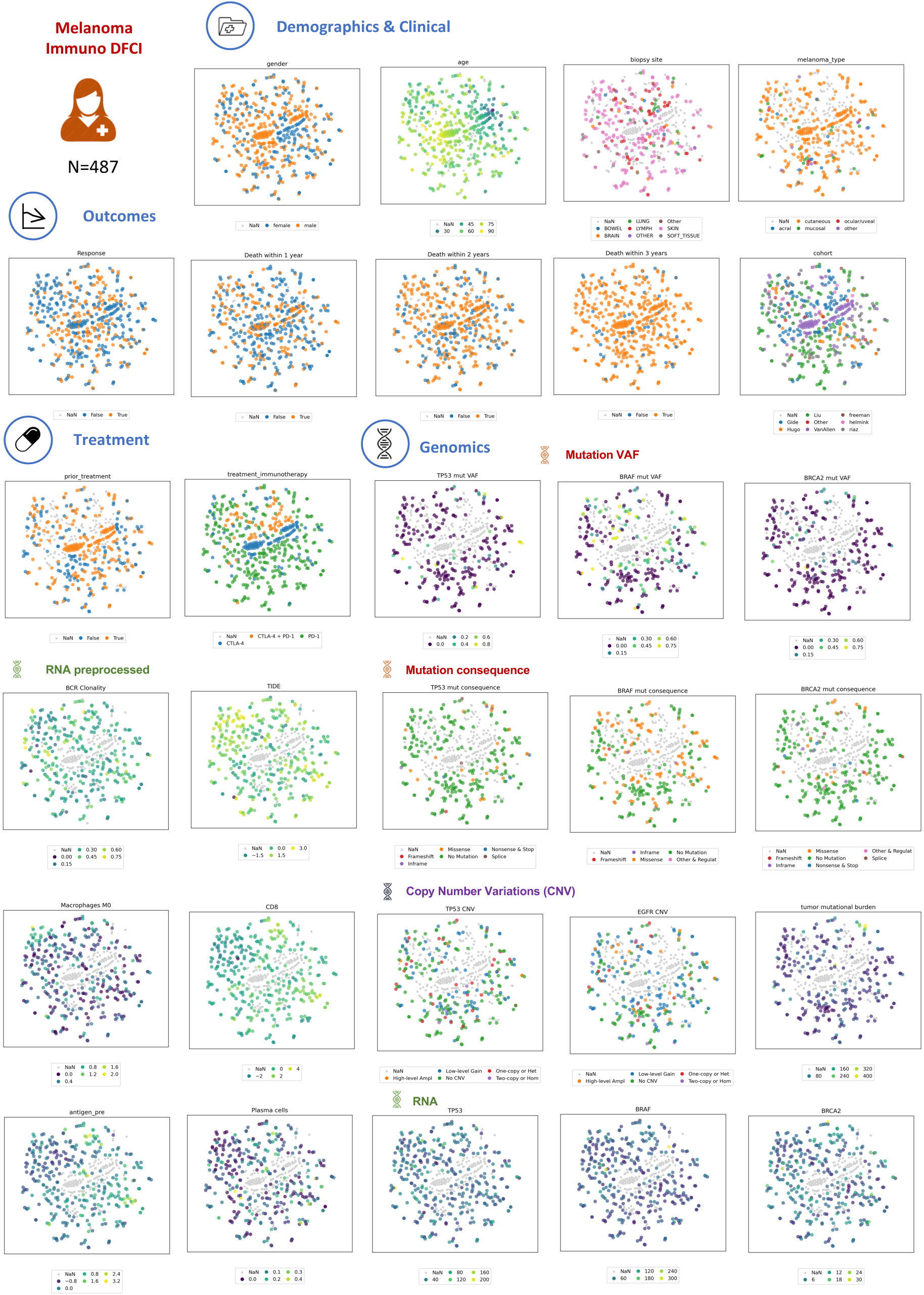
T-SNE Plots of the Melanoma Immuno 8RNAWES Cohort. Overview of all patients in the melanoma immune 8RNAWES cohort in the t-SNE space, colored by a subset of selected features. Cohort summary in Supplementary Table T6.

**Figure S7:**
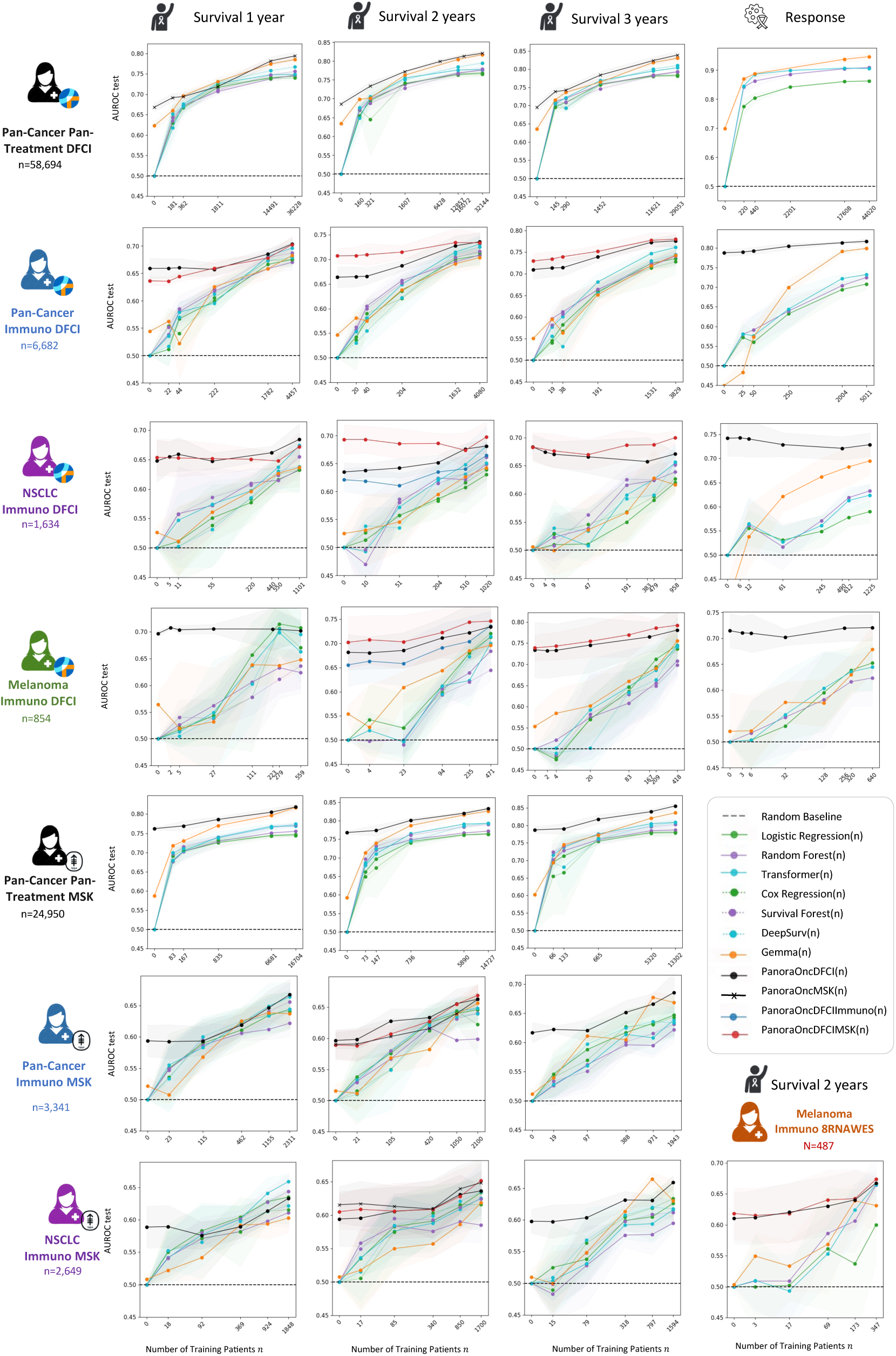
Extended Performance Results of *PanoraOncDFCI(n)* of Figure 2. We show extended performance results of *PanoraOncDFCI(n)* for the indicated cohorts per row and treatment outcomes per column.

**Figure S8:**
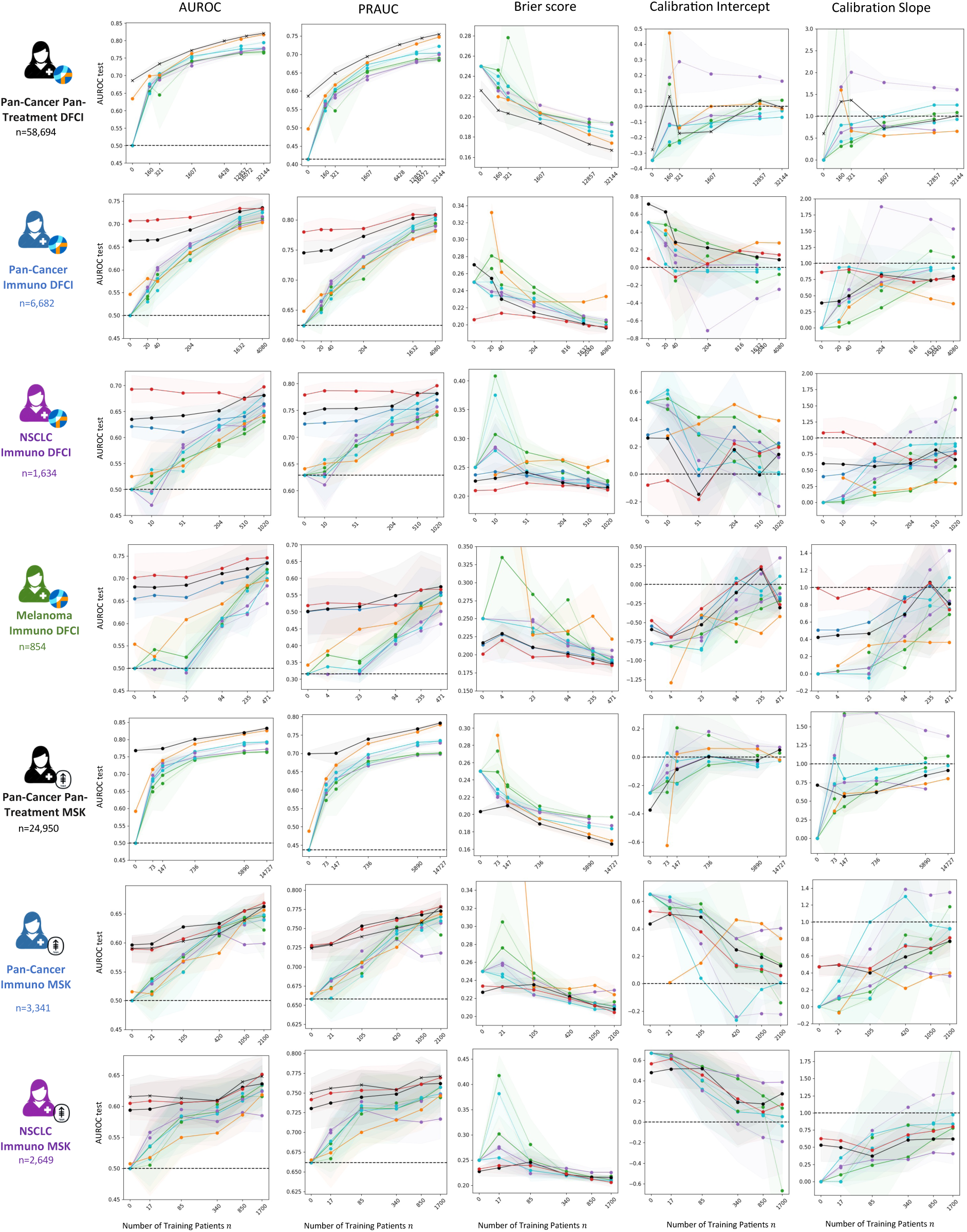
Extended Performance Results of Figure 3 for Different Metrics and Cohorts. Rows show the performance results for the indicated cohort, while rows compare different evaluation metrics.

**Figure S9:**
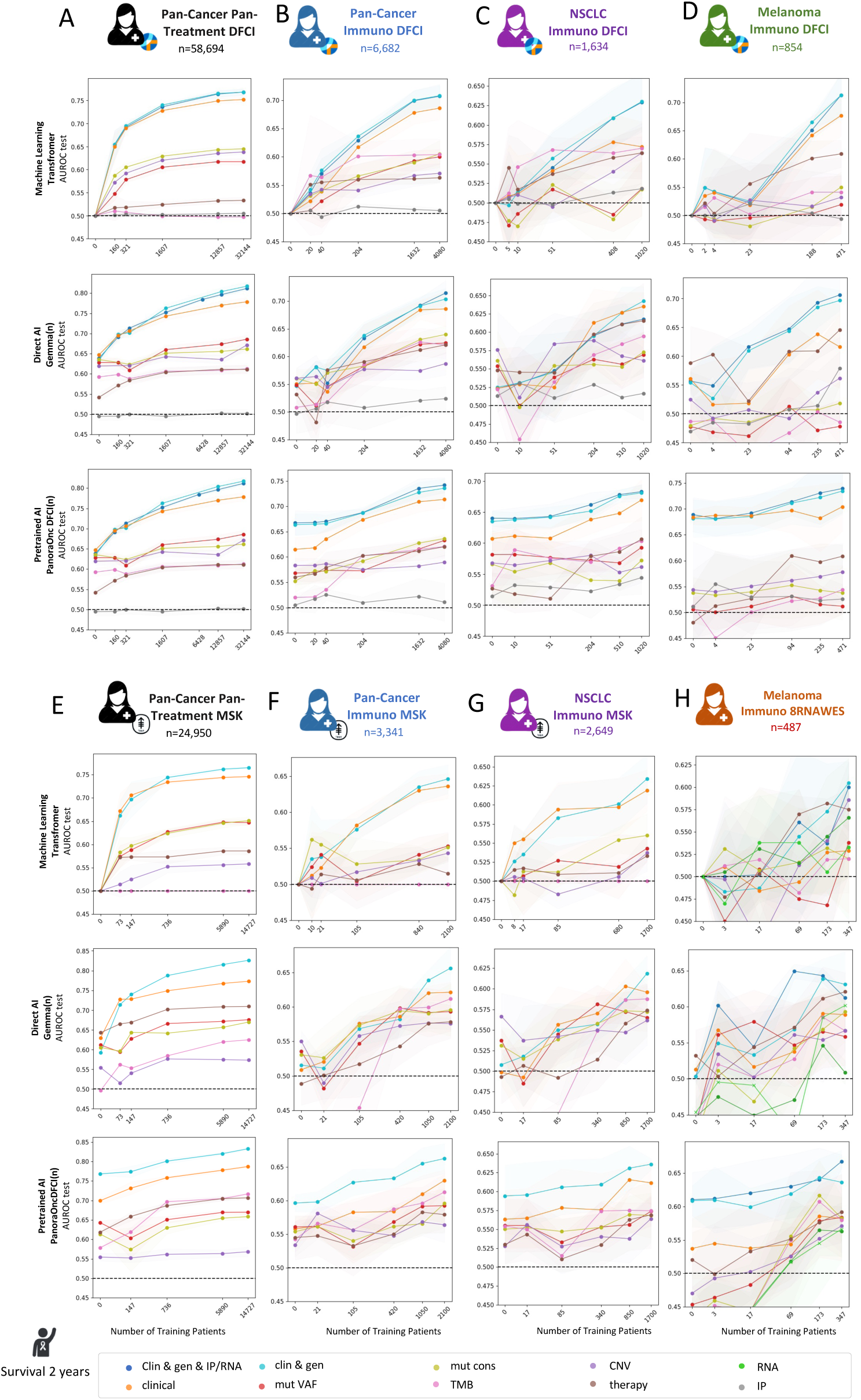
Extended Modality Results of Figure 3. Columns **(A-H)** show the performance results for the indicated cohort and different modalities, while rows compare direct ML (*Transformer*), direct AI (*Gemma*), and pretrained AI (*PanoraOncDFCI*).

**Figure S10:**
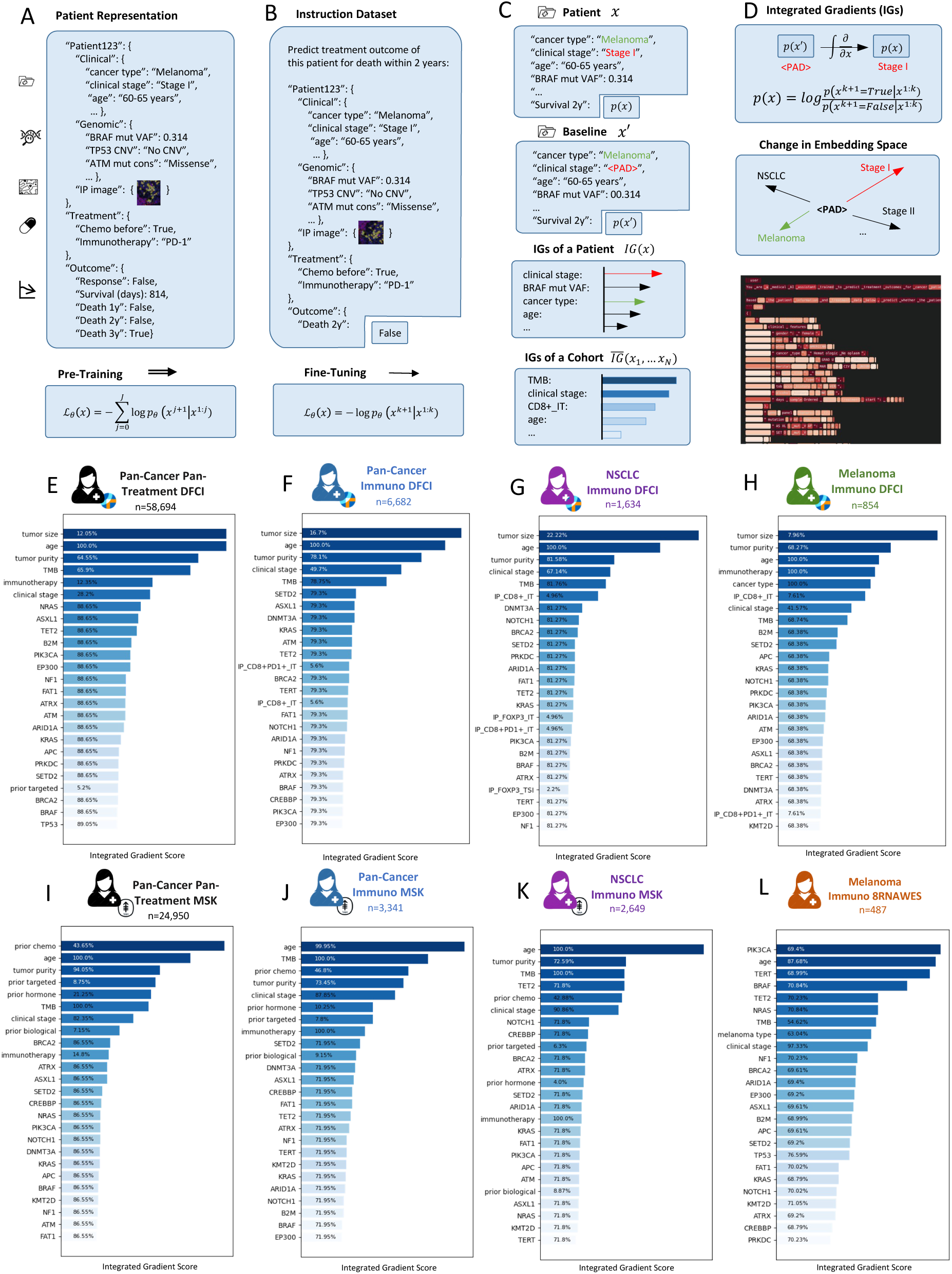
Patient Representations and Integrated Gradients. **(A-B)** Patient Representations used in PanoraOnc models. **(C-D)** Overview of the Integrated Gradient (IG) pipeline. **(E-L)** IG rankings of *PanoraOncDFCI* for the DFCI and MSK cohorts.

**Figure S11:**
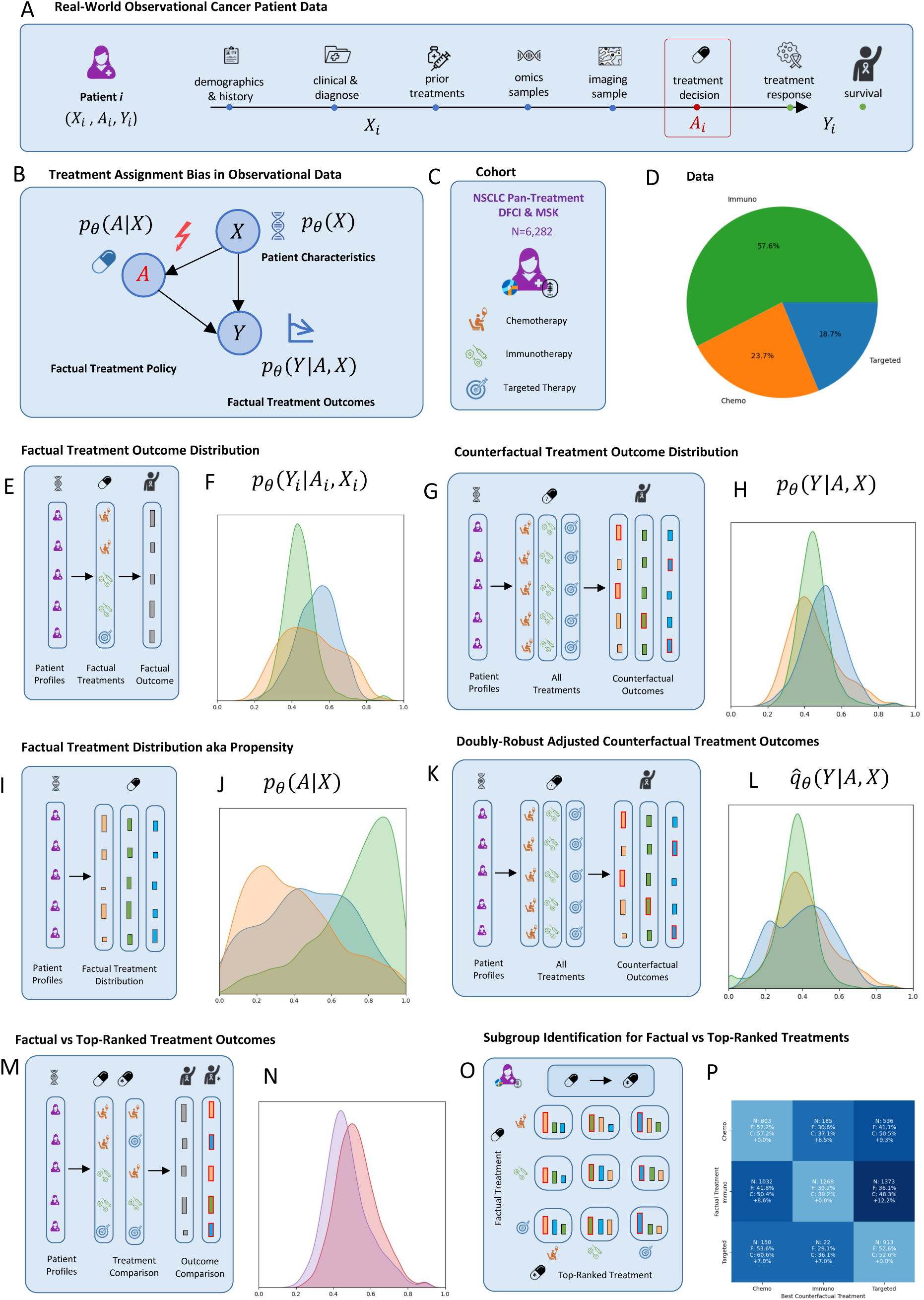
Bias-Aware Counterfactual AI Framework based on *PanoraOnc-PreDFCIMSK*. **(A)** Longitudinal multimodal patient representation, showing treatment decision at *t*_0_, patient features *X_i_* prior to treatment, treatment received *A_i_*, and observed outcomes *Y_i_*. **(B)** Schematic illustration of treatment assignment policies in observational real-world data, reflecting biases requiring adjustment. **(C)** Application to NSCLC cohorts from DFCI and MSK for chemotherapy, immunotherapy, and targeted therapy. **(D)** Factual treatment outcome distributions in the cohort, more details are provided in Fig. S13. **(E)** Model-predicted factual treatment outcomes. **(F)** Corresponding survival distributions in the NSCLC cohorts. **(G)** Model-predicted individualized counterfactual outcomes under all alternative treatments. **(H)** Corresponding counterfactual outcome distributions for all patients. **(I)** Model-predicted factual treatment distributions (propensity scores). **(J)** Corresponding distributions evaluated for the factual treatments in the cohort. **(K)** Schematic of bias-adjusted counterfactual outcomes obtained via TMLE. **(L)** Resulting bias-adjusted counterfactual treatment outcome distributions. **(M–N)** Comparison of factual versus top-ranked alternative treatments. **(O-P)** Stratification and identification of subcohorts that may benefit from alternative treatments using TMLE-adjusted predictions.

**Figure S12:**
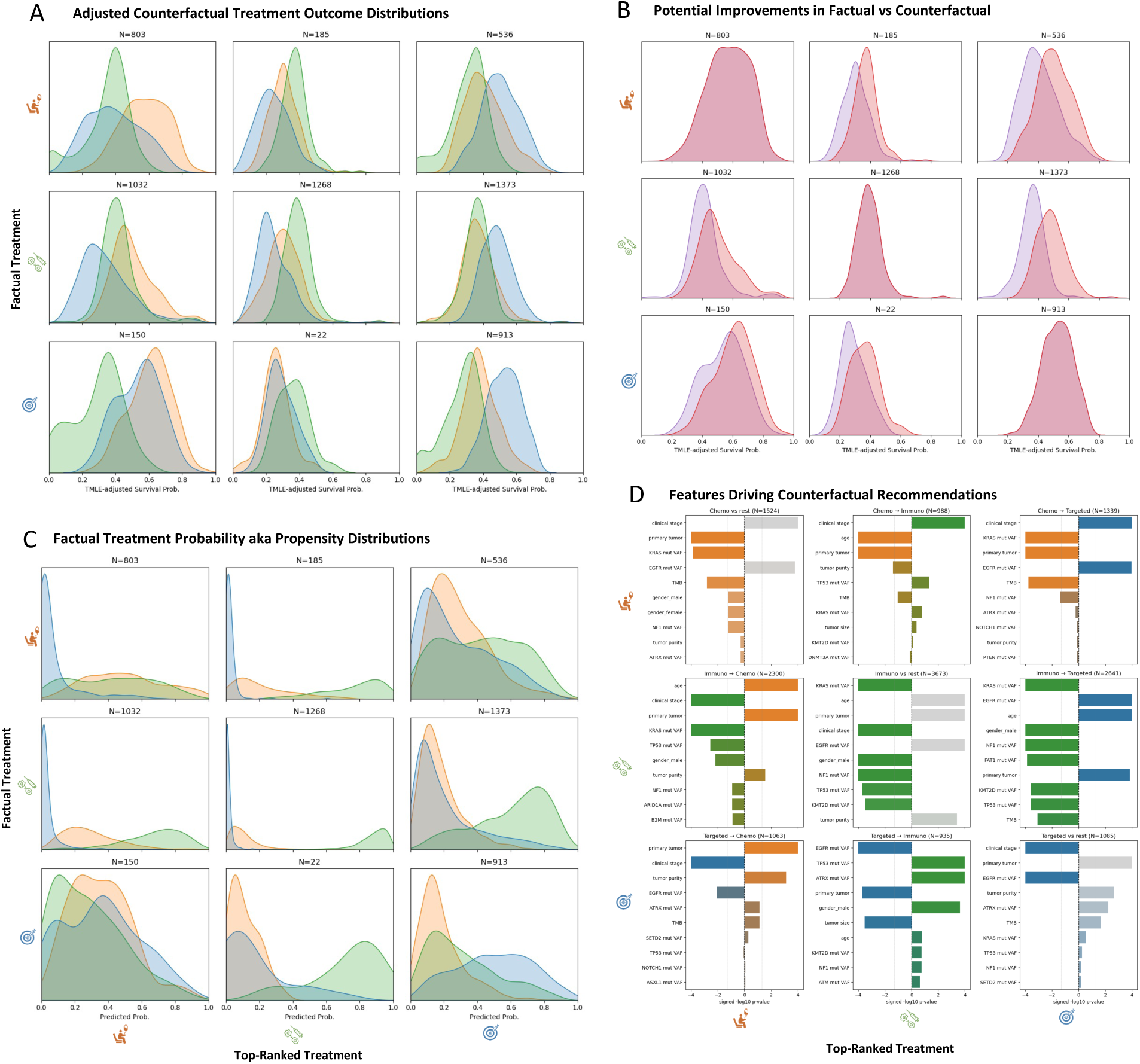
Factual-Counterfactual Treatment Analysis for the NSCLC Cohorts. **(A)** Bias-adjusted treatment outcome distributions from *PanoraOncDFCIMSK*, stratified by factual treatments (rows) and top-ranked counterfactual treatments (columns). **(B)** Factual versus top-ranked counterfactual treatment outcome distributions. **(C)** Estimated factual treatment probabilities within each subgroup, showing reasonable overlap across all factual-counterfactual comparisons (e.g., small probabilities for targeted therapy in the immunotherapy–chemotherapy subgroup are not consequential). **(D)** Top-ranked features for each subgroup, highlighting the clinico-genomic variables associated with the predicted switch versus stay outcomes.

**Figure S13:**
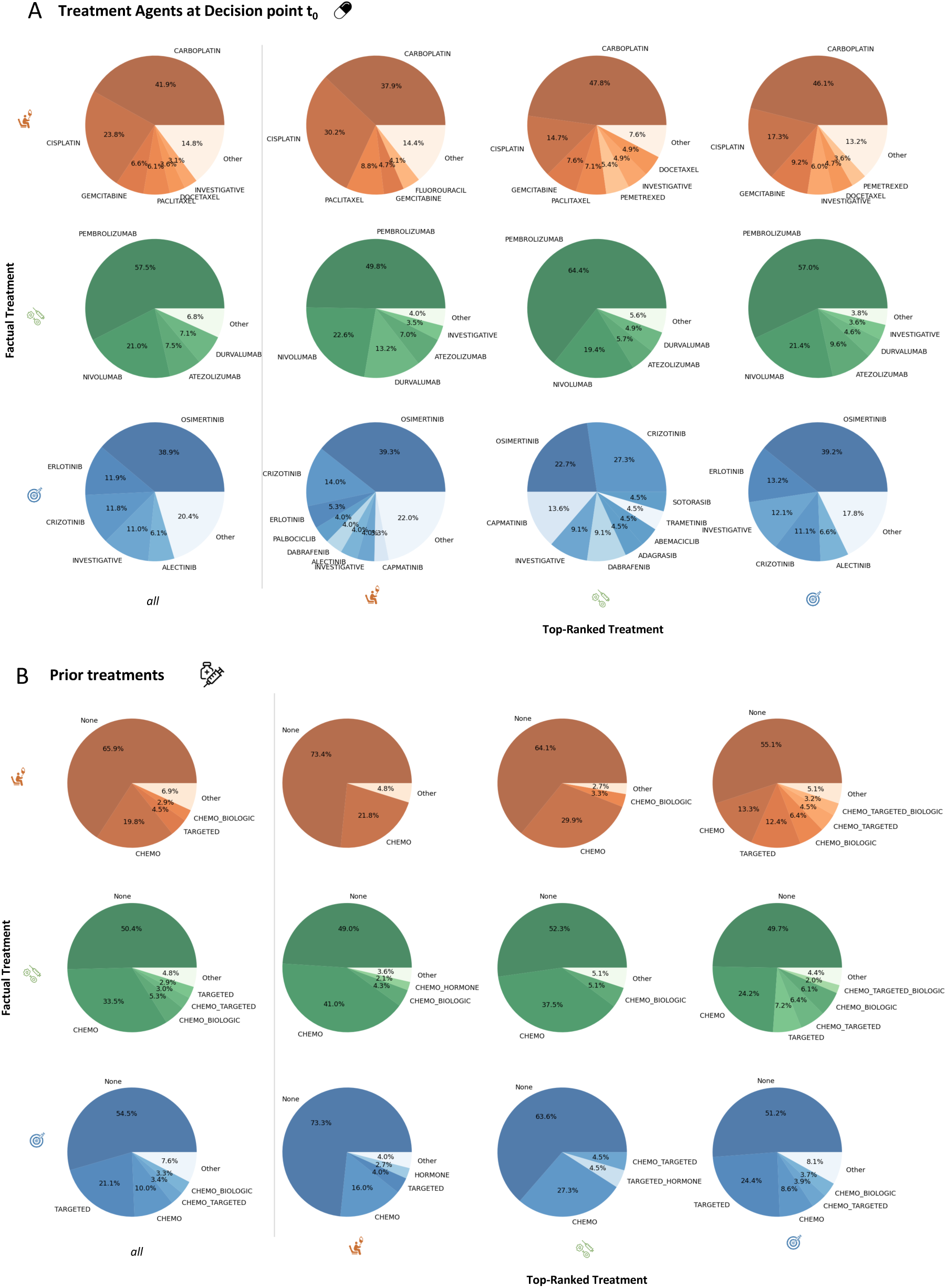
Treatment Distributions in Factual–Counterfactual Analysis. **(A)** Distribution of treatment agents at the treatment decision time point (t₀), stratified by factual treatments (rows) and top-ranked counterfactual treatment categories (columns); the first column shows the overall distribution across all counterfactuals. **(B)** Distribution of prior treatment categories, stratified analogously.

**Figure S14:**
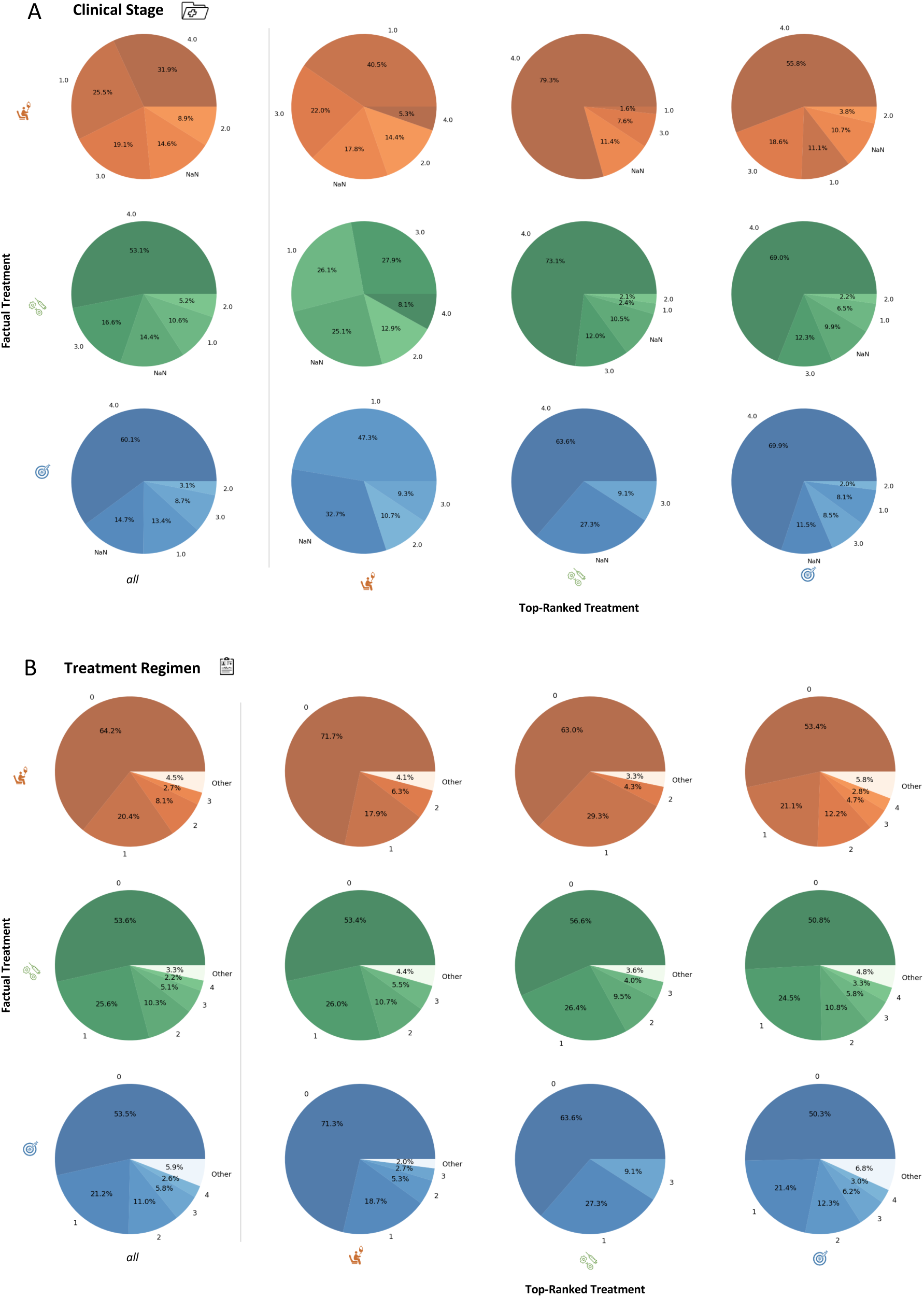
Stage and Regimen Distributions in Factual–Counterfactual Analysis. **(A)** Clinical stage and **(B)** treatment regimen distributions, stratified by factual treatments (rows) and top-ranked counterfactual treatment categories (columns), with the first column showing overall distributions.

**Figure S15:**
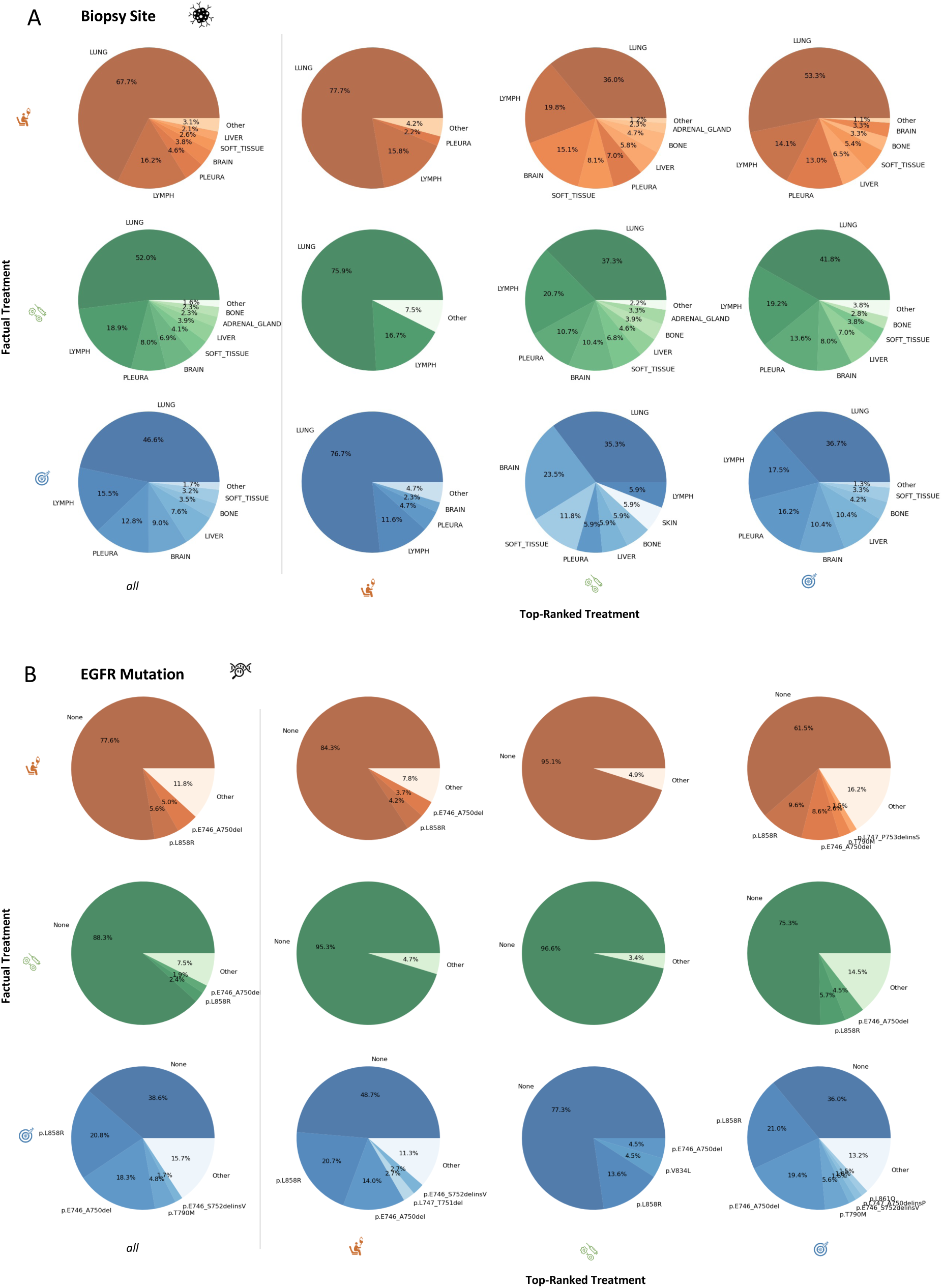
Biopsy Site and EGFR Mutation Distributions in Factual–Counterfactual Analysis. (A) Biopsy site and (B) EGFR mutation distributions, stratified by factual treatments (rows) and top-ranked counterfactual treatment categories (columns), with the first column showing overall distributions.

**Figure S16:**
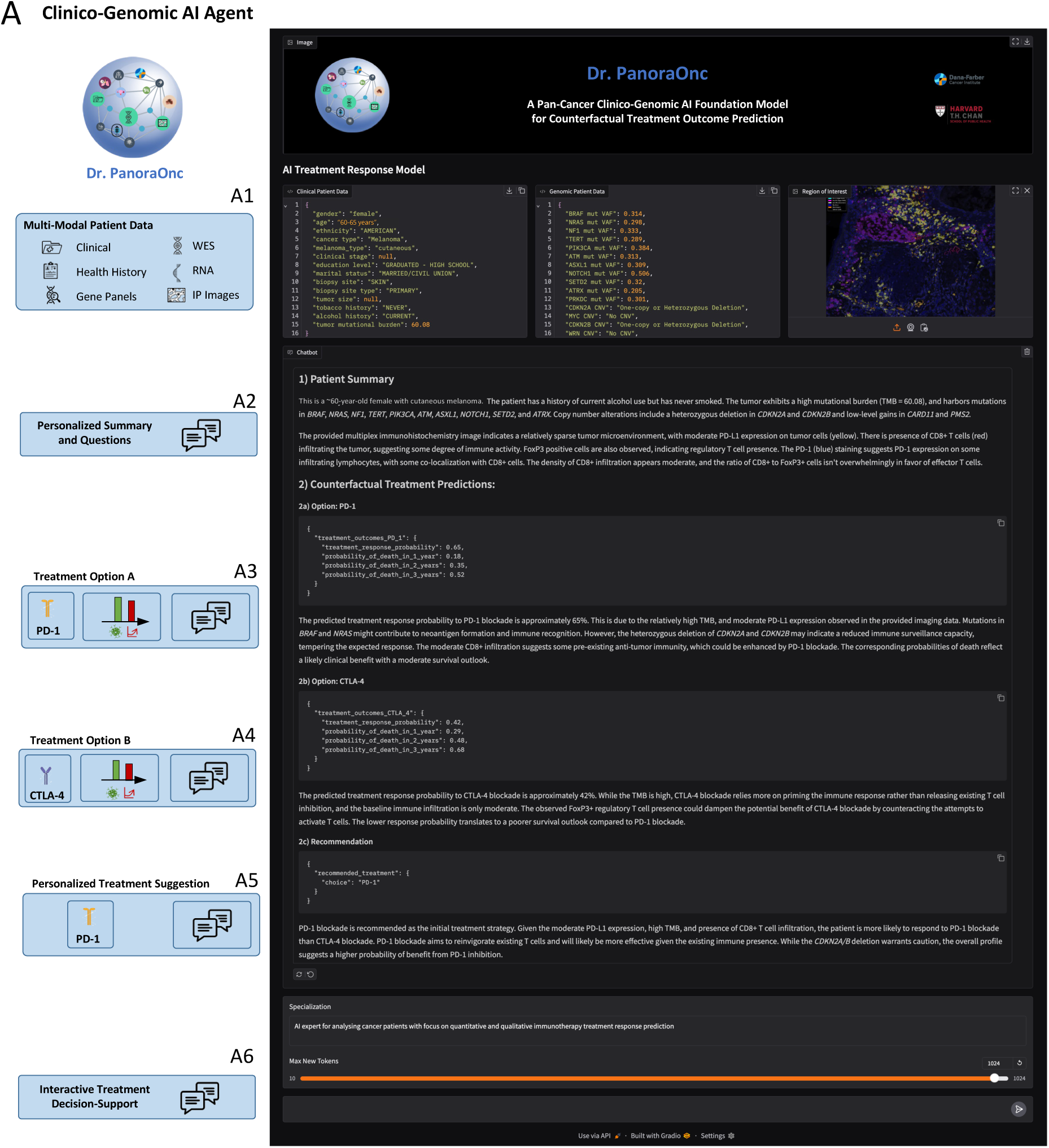
Interactive pan-cancer clinico-genomic AI agent for hypothetical treatment outcomes in personalized Oncology. **(A)** Illustrative patient case using the interactive AI agent, *Dr. PanoraOnc*, based on the pretrained *PanoraOncDFCI* VLM model. **(A1)** User-provided multimodal inputs, including clinical and genomic data, and a representative tumor image highlighting tumor–immune interactions. The AI module generates a patient summary **(A2)** with quantitative counterfactual predictions and reasoning for alternative treatment scenarios, illustrated for PD-1 **(A3)** and CTLA-4 blockade **(A4)**. The system integrates these results into personalized treatment suggestions **(A5)**. The interactive chat interface **(A6)** connects the AI model with patient data, images, and conversational history, enabling dynamic case summarization and data-driven and interactive therapeutic decision support. This system is a proof of concept only designed for research proposes to generate quantitative and interpretable hypothesis about treatment outcome scenarios and needs further rigorous external retro perspective, prospective and ultimately interventional evaluation.

## Notes

### Author Declarations

IRB approval was obtained for large-scale analysis of clinical and genomic features at Dana-Farber Cancer Institute, including treatment outcomes and NLP-derived response annotations, as well as for integrating multiplex immunofluorescence imaging data with clinical and genomic data. Controlled-access datasets, such as 8RNAWES, were obtained in accordance with repository-specific guidelines. Preprocessed and deidentified MSK clinical-genomic data are available without IRB requirements.

